# Predictors of Postviral Symptoms Following Epstein-Barr Virus-Associated Infectious Mononucleosis in Young People – Data from the IMMUC Study

**DOI:** 10.1101/2024.05.17.24307333

**Authors:** Maren Bodenhausen, Jonas Geisperger, Julia Lange de Luna, Johannes Wendl, Alexander Hapfelmeier, Lina Schulte-Hillen, Rafael Pricoco, Nina Körber, Tanja Bauer, Josef Mautner, Dieter Hoffmann, Peter Luppa, Silvia Egert-Schwender, Elfriede Nößner, Henri-Jacques Delecluse, Susanne Delecluse, Fabian Hauck, Christine Falk, Thomas Schulz, Marc-Matthias Steinborn, Andreas Bietenbeck, Alexandra Nieters, Lorenz Mihatsch, Katrin Gerrer, Uta Behrends, the IMMUC Study Group

**Author notes:** Corresponding author: Uta Behrends, M.D., Children’s Hospital, TUM School of Medicine and Health, Technical University of Munich, Koelner Platz 1, 80804 Munich, Germany. Bodenhauen M and Geisperger J contributed equally to this manuscript. Behrends U, Gerrer K, and Mihatsch L contributed equally to this manuscript. IMMUC Study Group team members are listed in the Acknowledgments.

## Abstract

**Background:** Epstein-Barr virus-associated Infectious Mononucleosis (EBV-IM) is a common disease following primary EBV infection in children and adolescents. While EBV-IM is mostly self-limiting, symptoms like fatigue may persist over several months or even result in myalgic encephalomyelitis/chronic fatigue syndrome (ME/CFS). This large clinical observational study aimed at identifying risk factors for protracted courses of EBV-IM in young people.

**Methods:** A cohort of N=200 children, adolescents, and young adults with acute primary EBV infection was recruited from hospitals and private practices. Data on the patients’ medical history as well as clinical and laboratory parameters were collected at a baseline visit (V1) within four weeks after symptom onset (T_onset_) and at two follow-up visits (V2 and V3) one and six months after T_onset_. Risk factors for protracted symptoms at V3 were modeled using multivariable logistic regressions.

**Results:** Protracted symptoms were observed in 55/183 (30.1%) and protracted fatigue in 34/181 (18.8%) patients at V3. A medical history indicating an increased susceptibility to infectious diseases as well as distinct severe IM symptoms, e.g. severe gastrointestinal symptoms, were significantly associated with protracted disease [OR: 2.31; P=0.011 and OR: 3.42; P=0.027] and with chronic fatigue [OR: 2.98; P=0.006 and OR: 3.54; P=0.034], respectively. Occurrence of twelve or more clinical and laboratory parameters until and including V1 discriminated between fatigue and no fatigue at V3 [OR 2.43, P=0.033].

**Conclusion:** A clinical history of immune dysregulation as well as distinct severe IM symptoms might predict protracted post-viral disease and thus help in the identification of young patients at risk.

**Highlights:**

- Severe gastrointestinal symptoms are associated with protracted course of Epstein-Barr virus-associated Infectious Mononucleosis (EBV-IM).
- Signs of immune dysregulation prior to EBV-IM can indicate an increased risk of protracted symptoms.
- Greater number of initial symptoms helps to identify patients developing postviral fatigue.

## INTRODUCTION

Infectious mononucleosis (IM) develops in up to 75% of adolescents and young adults after primary infection with Epstein-Barr virus (EBV) [1]. Symptoms typically include fever, lymphadenopathy, tonsillopharyngitis, splenomegaly, and fatigue, and usually are self-limiting within two weeks to three months [1]. However, acute complications such as kissing tonsils, splenic rupture, or life-threatening hemophagocytic lymphohistiocytosis (HLH) [1, 2] can occur, and lymphoproliferative disease can manifest in immunocompromised patients [3]. Moreover, post-viral symptoms may persist for weeks or years and may fulfill the clinical criteria of myalgic encephalomyelitis/chronic fatigue syndrome (ME/CFS) [4, 5], significantly compromising daily activities [6].

Risk factors for long-term IM sequelae are still under investigation, and various scores have been published to assess IM severity, complexity, and protraction [7-13]. In a large cohort of college students, physical symptoms and immune dysregulation have been identified as risk factors for ME/CFS following IM [14], and autonomic symptoms along with distinct immune markers were suggested as predictors of severe ME/CFS [15].

Here, we report on a novel IM scoring system and on candidate risk factors for protracted symptoms identified in the Munich Infectious Mononucleosis (IMMUC) study. This observational, prospective, longitudinal study investigates a comprehensive set of routine and experimental data in 200 children, adolescents, and young adults with recent IM to identify biomarkers and causative factors of complicated disease trajectories, including chronic fatigue.

## MATERIAL & METHODS

### Ethics, Study Setting, and Inclusion Criteria

The IMMUC study was approved by the TUM University Hospital Ethics Committee (No. 112/14) and adheres to the 1964 Declaration of Helsinki, its later amendments, and the ICH-GCP regulations. All participants and/or legal guardians gave written informed consent. The study was registered at ClinicalTrials.gov (<u>NCT06002802)</u>. 200 participants with recent IM onset were prospectively recruited from hospitals or private practices in or around Munich between February 2016 and June 2019 and observed up to 12 months after IM onset. General, clinical, and virological inclusion criteria are summarized in **Tab. 1**.

**Table 1.** IMMUC Inclusion Criteria.

| A) General Criteria |  |
| --- | --- |
| I | Age ≤ 39 years |
| II | No pregnancy ≤ 12 months prior to recruitment |
| III | Written informed consent |
| B) Clinical IM Criteria |  |
| At least 1 of the following IM symptoms with new onset ≤ 28 days prior to recruitment | |
| I | Tonsillopharyngitis |
| II | Lymphadenopathy |
| III | Fever ≥ 38.5°C |
| IV | Fatigue |
| C) Virological IM Criteria |  |
| At least 1 of I.1-4 at V1 <u>and</u> II at V1 <u>and</u> III at V1 or Tonset <u>and</u> IV |  |
| I | 1. EBV DNA positive in cell fraction<br>2. EBV DNA positive in plasma<br>3. EBV VCA IgM positive in any test<br>4. EBV VCA IgG positive in any test |
| II | EBNA-1 IgG maximal 1+ in immunoblot |
| III | EBNA-1 IgG negative in any test |
| IV | All data Tonset-V3 support EBV primary infection |

### Study Visits, Study Periods and Study Data

Patients were seen at visit 1 (V1) within 28 days after onset (DAO) of IM (DAO 0=T_onset_). V2 was scheduled at four to six weeks after T_onset_ and V3 at four to 12 months after T_onset_. A defined set of virological (IM_Vir_), clinical chemistry (IM_Lab_), and clinical (IM_Clin_) features of IM was investigated at each visit. Additionally, medical history data and external laboratory data were collected for the time periods before T_onset_ and between T_onset_, V1, and V2, to generate combined datasets for the history period (HP) before T_onset_, the very early period (VEP) from T_onset_ up to and including V1, and the early period (EP) from T_onset_ up to and including V2 (**Fig. 1**). All visits included a structured interview, a physical examination, and peripheral blood collection for routine (reported here) and experimental (reported elsewhere) laboratory analyses. Splenic sonography was performed at V1 and V2 and in case of persistent splenomegaly at V3. For a detailed account of the collected study data see **Suppl. Methods, Suppl. Material M1,** and **Suppl. Material M2.**

**Figure 1.**
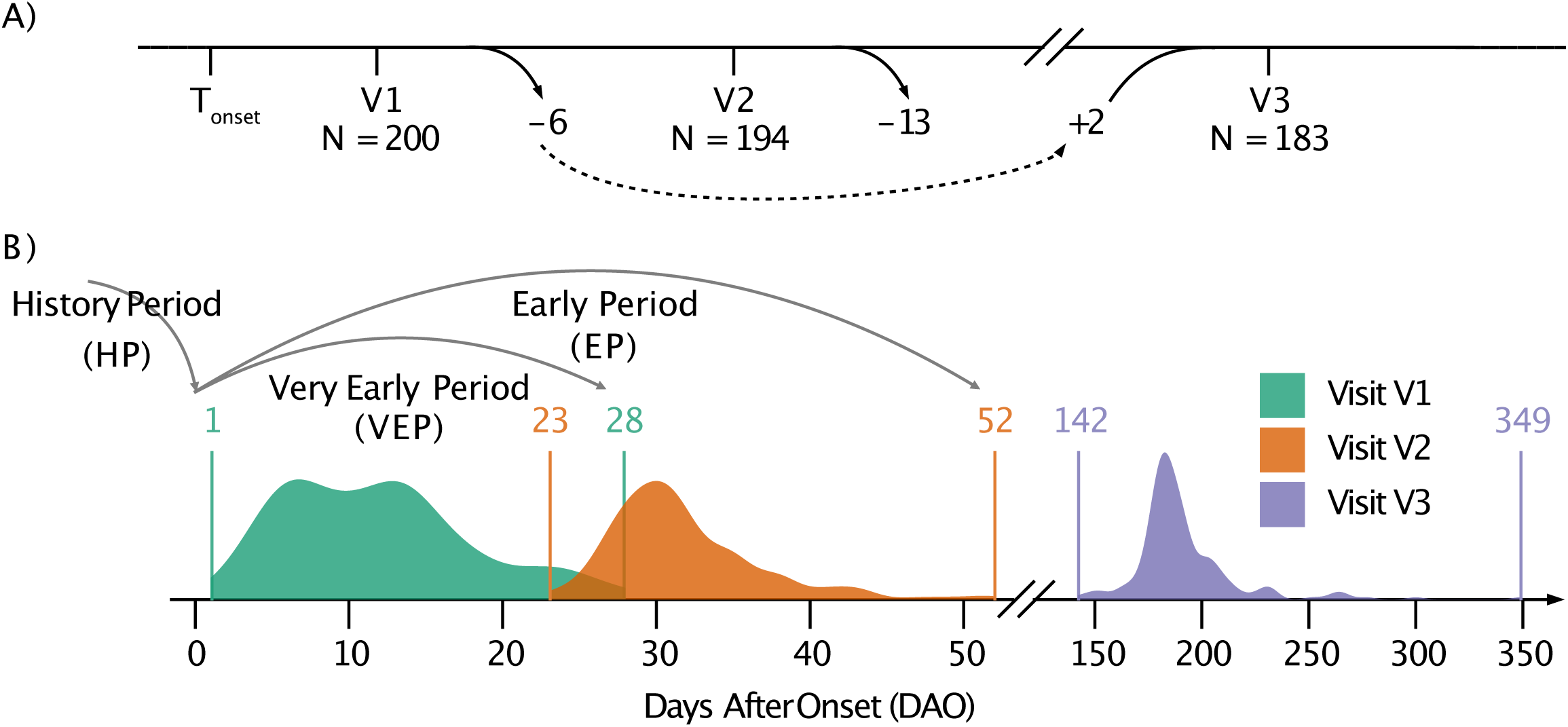
Study Setting with Number of Participants (A), Study Visits, and Study Periods (B). (A) Two hundred patients were recruited, of which 194 were seen at visit 2 (V2) and 183 at visit 3 (V3), with two patients not seen at V2 but again at V3 and another four dropping out before V2. (B) The X-axis depicts the days after onset (DAO) of IM from day 0 up to day 349 when the last patient was seen for V3. V1 was scheduled up to 28 DAO, V2 23-52 DAO, and V3 142-349 DAO. Time periods were defined as medical history period (HP) as the time period before T_onset_, i.e., 0 DAO, the very early period (VEP) from IM onset (T_onset_) until and including V1 and the early period (EP) from T_onset_ until and including V2.

### IM Scoring

A score was developed to grade severity (Smax), complexity (Cmax), and protraction of individual IM features and overall IM at each visit and for each period (**Tab. 2**). Up to five grades of severity (S0-5) were evaluated for each IM feature, and the highest severity grade of all distinct features was used to grade the overall IM (**Suppl. Mat. M2**). By calculating the number of IM features, complexity grades were determined for the overall IM (C0-C23), IM_Clin_ (C0-15), and IM_Lab_ (C0-8) IM. Protraction grades were defined as P0 (feature/IM not present anymore at V2 and V3), P1 (feature/IM present at V2 but not at V3), or P2 (feature/IM present at V3). Since the clinical significance of mild lymphadenopathy and splenomegaly at V3 was unclear, only S2-4 of the latter at V3 were counted as P2. The grading of fatigue at V3 differed from that at V1 and V2 due to an increasing clinical impact of fatigue over time.

**Table 2.**
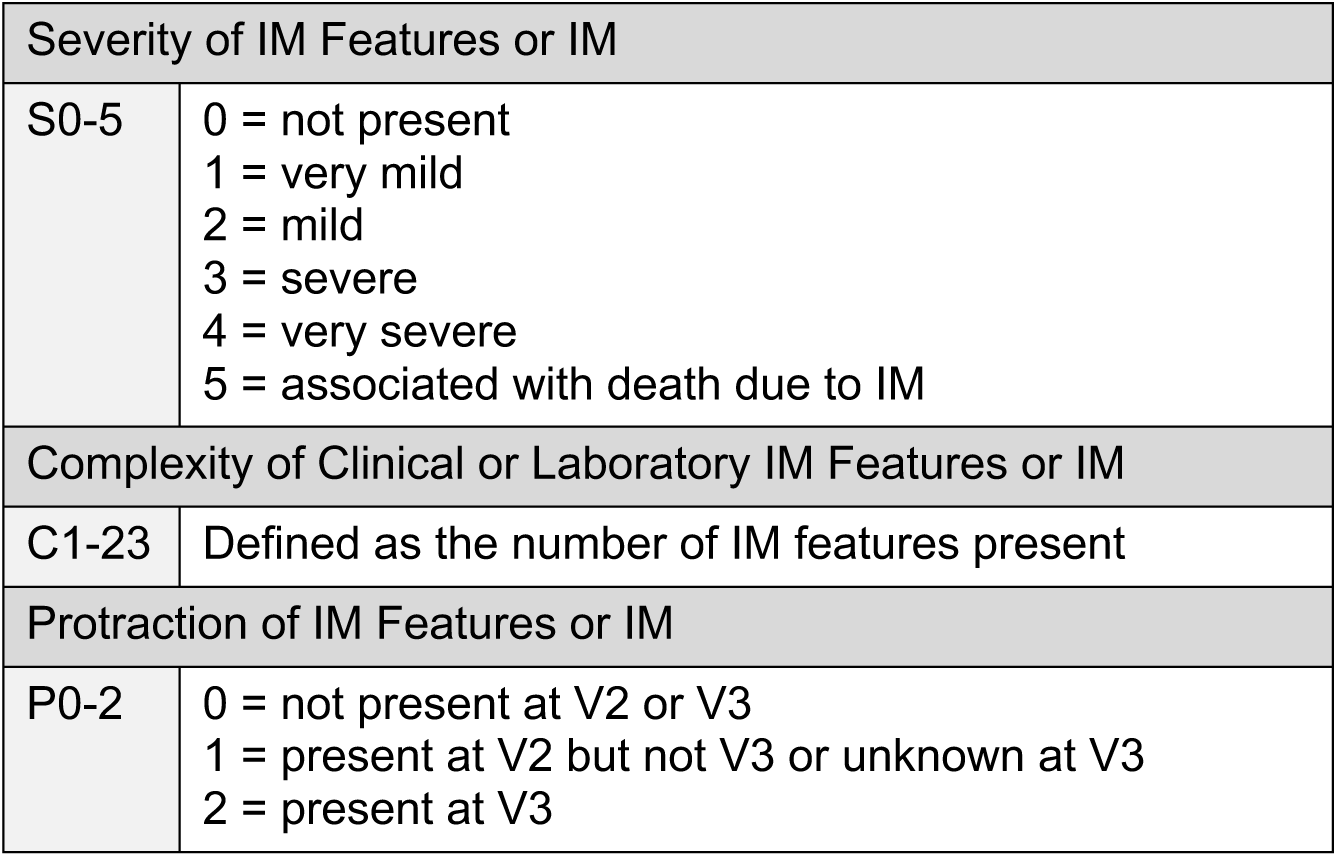
IMMUC Scores.

### Outcome Measures, Predictor Variables in Multivariable Logistic Regression Models

Long-term IM protraction (P2-IM vs. P0/1-IM) or chronic fatigue (P2-fatigue vs. P0/1-fatigue) were the binary outcome measures to be modelled. Predictor variables were either measured repetitively at visits V1 and V2 (quantitative and categorized IM_Vir_, IM_Lab_ and IM_Clin_ features) and thus, included into the *visit-dependent* analyses (see below) or only once during the periods HP, VEP, and/or EP (quantitative and categorized baseline patient characteristics as well as Cmax-IM, -IM_Lab_, -IM_Clin_, Smax-IM, -IM_Lab_, and -IM_Clin_ features) and thus included into the *period-dependent* analyses (see below).

### Sample Size Calculation

We expected approximately 10% of patients with P2-IM, and therefore, set the sample size to N=200 patients (**Suppl. Methods)**.

### Visit-Dependent Analyses

Each multivariable logistic regression model included one of the predictor variables measured at V1 and V2, and sex and age as independent variables. Since individual patients were measured repetitively, an interaction effect between the predictor variable and DAO was included in each model. The effect of a predictor variable can thus be described in dependence on DAO. These models also determined a time interval at which the OR of the predictor variable was significantly different from 1. The robust Huber-White estimator accounted for minor misspecifications of the model assumption due to repeated measurements.

### Period-Dependent Analyses

Each multivariable logistic regression model included one of the predictor variables measured once for either of the three periods: HP, VEP, or EP and sex and age as independent variables. A grid search was used to find the optimal cut-off value for Cmax-IM and Cmax-IM_clin_ in terms of the area under the curve (AUC).

For the *visit-* and *period-*dependent analyses, models with less than five observations per independent variable per category variable of the outcome variable were discarded to prevent overfitting. Only significant results are presented here due to the abundance of variables investigated.

### Other Statistical Analyses

Hypothesis tests were performed at exploratory 5% significance levels. P-values are exploratory and thus not corrected for multiple testing. Analyses were performed using R, version 4.1.3 (The R Foundation for Statistical Computing, Vienna, Austria) [16].

## RESULTS

Of 200 study participants assessed at V1, 194 and 183 attended V2 and V3, respectively (**Fig. 1A**). Study visits (V1, V2, V3) and study periods (HP, VEP, EP) are depicted in **Fig. 1B**.

### Baseline Patient Characteristics

Detailed data are shown in **Suppl. Table S1A to S1F.** The cohort included 92 children (1-11 years), 84 adolescents (12-17 years), and 24 adults (18-33 years). 54.5% patients were female. The mean age was 11.99±6.08 years (range: 1–33) (male: 11.76±6.89, female: 12.18±5.33; t-test: P=0.632). A history of increased susceptibility to infections and/or immune dysregulation (ELVIS and/or GARFIELD criteria [17]) prior to IM was reported by 47.5% of patients (**Suppl. Table 1E)**. When asked for any fever or antibiotic therapy during the six months prior to IM, 37.0% and 13.6% of patients provided positive answers, respectively. 35.7% and 6.0% of patients remembered ≥1 or ≥2 family members with a history of IM.

### Virological Features of IM

Detailed data are provided in **Fig. 2**.

**Figure 2.**
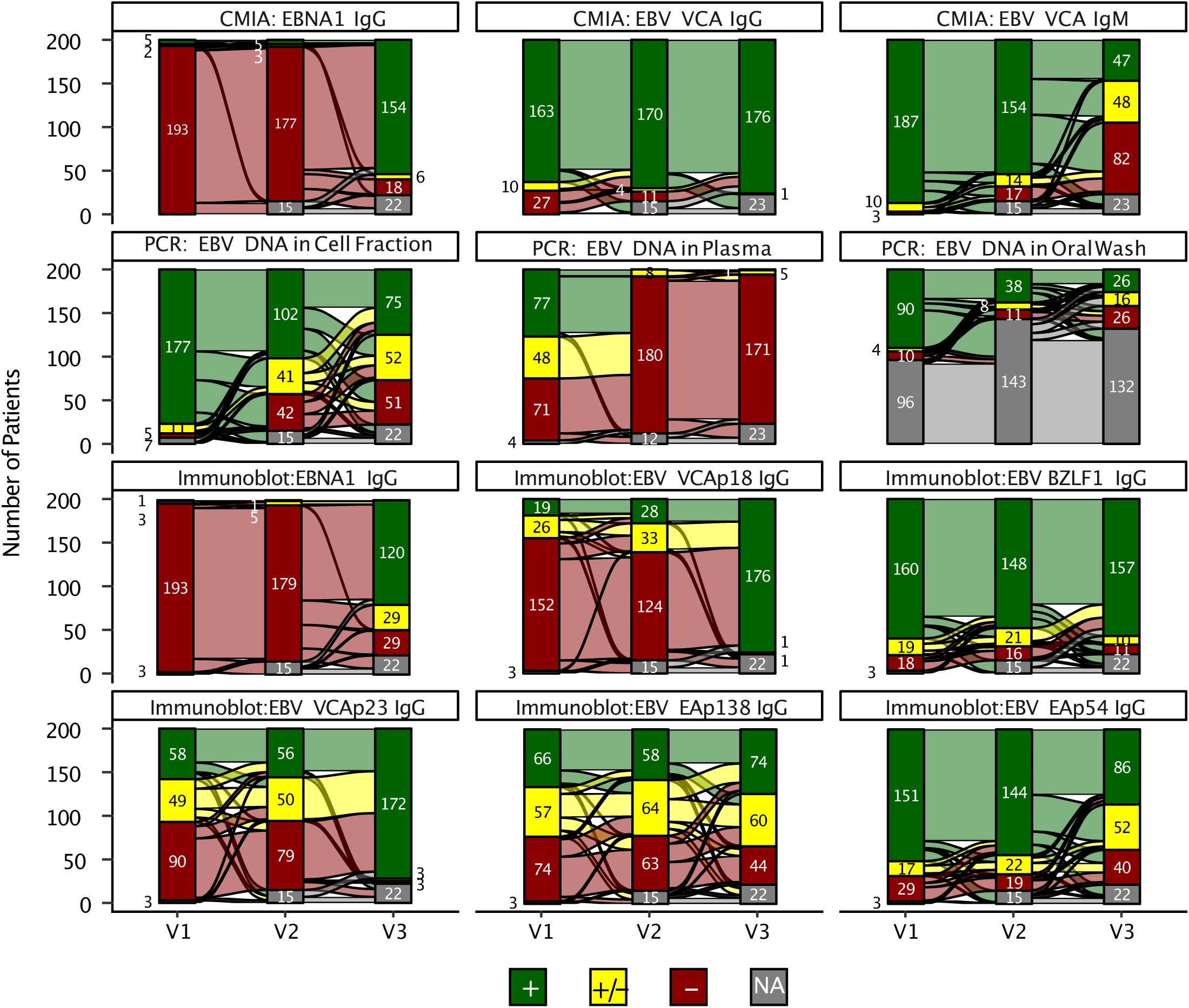
Virological IM Features. Alluvial plots of virological features of IM (IM_Vir_). The first row shows the results of the chemiluminescent microparticle immunoassay (CMIA) for EBNA1 IgG, EBV VCA IgG, and EBV VCA IgM. The second row shows the PCR results for detecting EBV DNA in cell fraction, in plasma, and in oral washes. The third and fourth rows show the Immunoblot results for EBNA1 IgG, EBV VCAp18 IgG, EBV BZLF1 IgG, EBV VCAp23 IgG, EBV EAp138 IgG, and EBV EAp54 IgG. Red depicts a negative result, yellow is an intermediate, and green is a positive result of the respective method. Grey indicates missing data. The bands between the stacked columns show the development of the test results. CMIA at V1, V2, and V3 detected VCA IgM in 93.5%, 83.2%, and 26.6%, VCA IgG in 81.5%, 91.9%, and 99.4%, and EBNA-1 IgG in 2.5%, 2.7% and 86.5% cases, respectively, indicating negative VCA IgM and/or IgG at V1, negative EBNA-1 IgG at V3, and persisting VCA IgM at V3 in some cases as expected. Immunoblots at V1, V2, and V3 showed IgG against VCAp18 in 9.6%, 15.1% and 98.9% against EAp54 in 76.6%, 77.8% and 48.3%, against EAp138 in 33.5%, 31.4% and 41.6% patients, and against at least one EA in 80,7% 81,6% and 60,7% respectively, demonstrating early VCAp18 IgG and late EA IgG in a significant number of patients. PCR at V1, V2, and V3 revealed EBV_oral_ in 86.5%, 66.7% and 38.2%, EBV_cells_ in 91.8%, 55.1% and 42.1% and EBV_plasma_ in 39.3%, 0.0% and 0.6% patients, respectively, with detectable EBV DNA in recovered cases as expected. The highest viral load at V1, V2, and V3 were 38,100.0 Geq/10^5^, 8,190.0 Geq/10^5^, and 2,330.0 Geq/10^5^ for EBV_cells_, and 130,000.0 IU/ml, 0 IU/ml and 530.0 IU/ml for EBV_plasma_, indicating that very high viral load in peripheral blood is short-lived as described [8].

### Severity, Complexity, and Protraction of IM

Comprehensive data on IM severity, complexity, and protraction are provided in **Fig. 3A-C**, **Suppl. Fig. S1, Suppl. Fig. S2** and **Suppl. Tab. S2**. IM severity and complexity decreased from V1 to V3 as expected. At least one IM feature persisted in 52.3% at V2 (P1) and in 27.9 % at V3 (P2) (**Fig. 3C**). 34 (18.8%) patients suffered from fatigue at V3, with mild fatigue (S1) reported by 82.4% and moderate fatigue (S2) (“reduced education/work/club sports”) by 17.6% of these patients (**Fig. 3A,C**).

**Figure 3.**
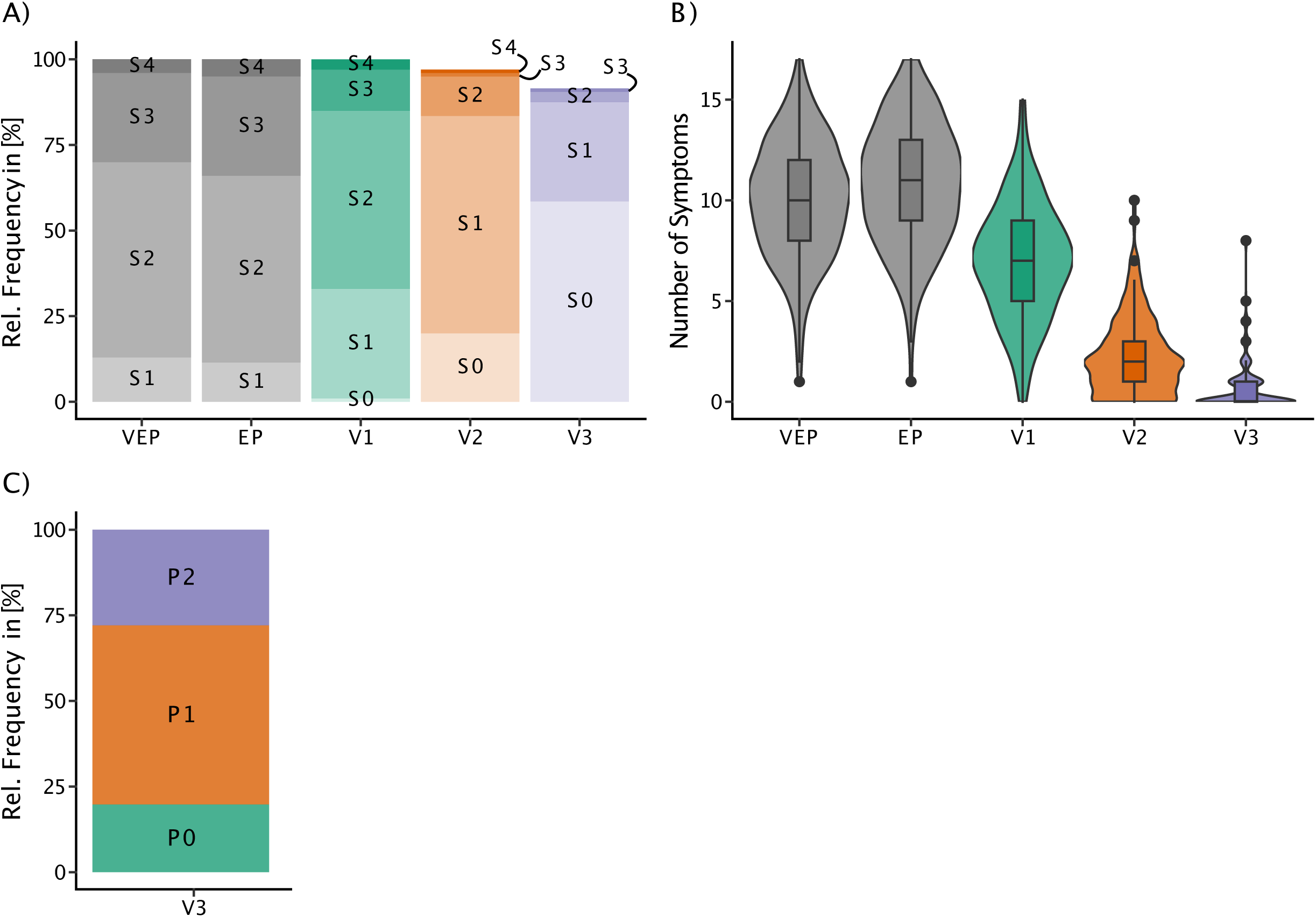
Main Clinical Outcomes. Summary of outcomes measured for the very early period (VEP, grey), the early period (EP, grey), and at the visits 1 – 3 (colored). The severity of all symptoms, according to the IMMUC score (A), includes clinical and laboratory symptoms. The violin plot of the complexity score (B) shows the number of observed clinical and laboratory symptoms. Protraction (C) indicates if patients had symptoms until and including V1 (P0), V2 (P1), or V3 (P2).

### Predictors for Long-Term Protraction (P2-IM)

Significant predictors of any IM feature at V3 (P2-IM) of the *visit-*dependent analyses are shown in **Fig. 4A**. Early positivity for VCAp18 IgG was the only risk factor, with decreasing effect over time (negative slope in **Fig. 4A**). Protective effects were also observed for early atypical lymphocytosis (until 6 DAO), tonsillopharyngitis (until 9 DAO) and leukocytosis (until 4 DAO). Complexity of IM and IM_Clin_ were significant risk factors for P2-IM from DAO=6 and DAO=5 onwards, respectively. Additional IM_Clin_ features predicted P2-IM, again with a delay of few days after T_onset_ and depending on severity (positive slope in **Fig. 4A**). Considering the AUC, the best predictor variables were complexity of IM_Clin_ (AUC 0.68), complexity of IM (AUC 0.66) and presence of other airway symptoms [S1-4 vs. S0] (AUC 0.62).

**Figure 4.**
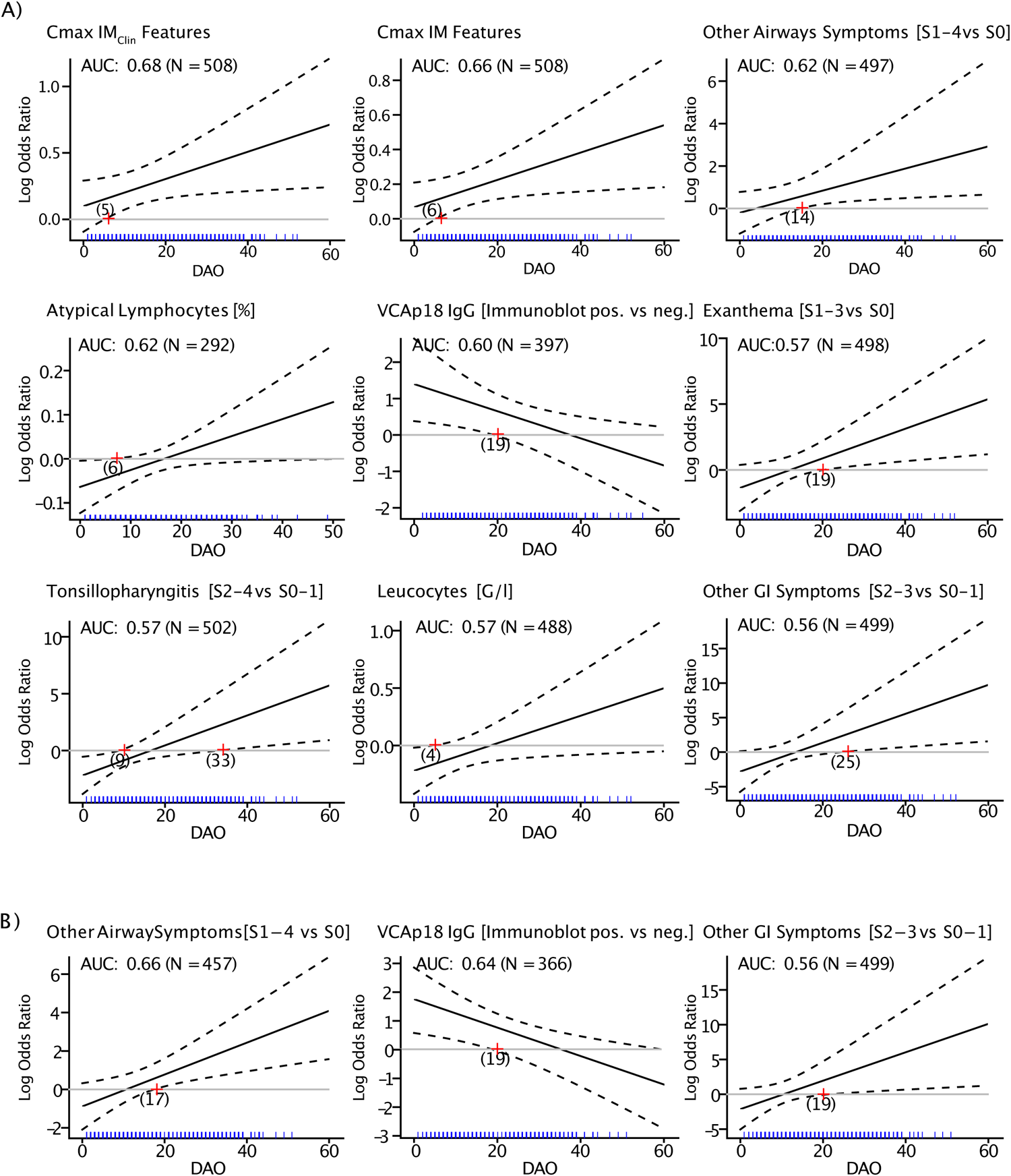
Results of the Visit-Dependent Analyses. Multivariable logistic regression models were used to model the effect of each visit-dependent variable for protraction (P2-IM) (A) and fatigue at V3 (P2-fatigue) (B). A separate model was estimated for each predictor variable, with sex and age as additional covariables. Each subplot shows how the log OR (black line) changes depending on the days after the onset (DAO) of IM symptoms. The dashed lines show the pointwise 95%-confidence band. A log OR of 0 is equivalent to an OR of 1. Thus, whenever the zero line (grey) is not included in the 95%-confidence band, the OR is significantly different from 1. The DAO where the zero line intersects the boundary of the 95%-confidence band is marked by a red cross, with the respective DAO in parenthesis. The blue rug shows when the respective (multiple) measurements were taken. The respective AUC of each model and the number of measurements included in the model is shown in the top left corner of each plot.

Significant predictors of any IM feature at V3 (P2-IM) of the *period-*dependent analyses are shown in **Fig. 5A**. IM_Clin_ complexity in the VEP and EP were the best predictors in terms of the AUC. The ROC analysis indicated that a cut-off of ≥10 and ≥9 IM_Clin_ features during the VEP (OR 2.61, 95%-CI: [1.27, 5.37]) and EP (OR 2.63, 95%-CI: [1.37, 5.03]) provided the greatest AUCs for predicting P2-IM, respectively. Analogously, a cut-off of ≥12 and ≥13 IM features provided the greatest AUCs for the VEP (OR 2.47, 95%-CI: [1.26, 4.87]) and EP (OR 2.99, 95%-CI: [1.49, 5.98]), respectively. Interestingly, ≥1 positive ELVIS criterion was also identified as a significant risk factor for P2-IM.

**Figure 5.**
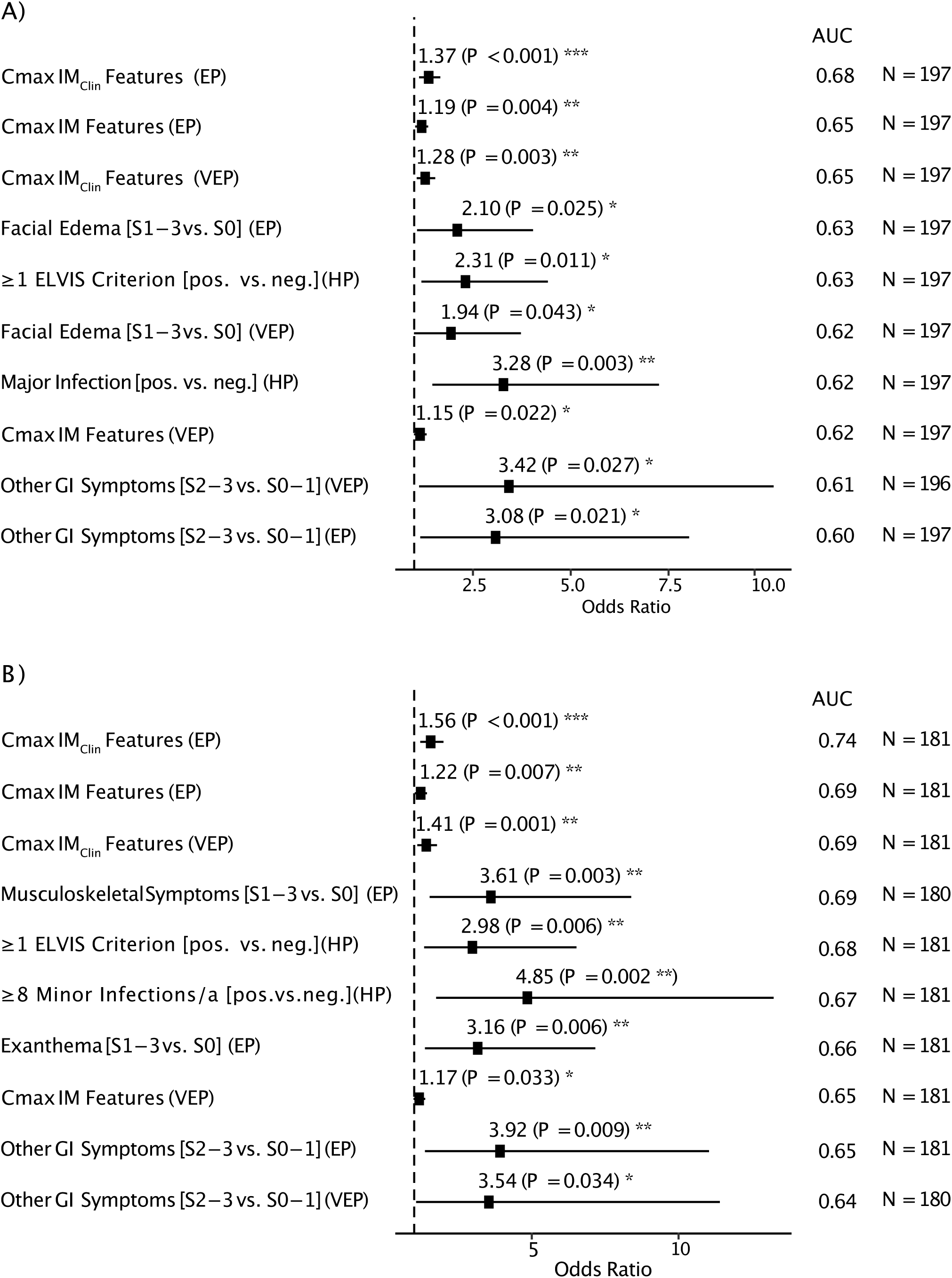
Results of the Period-Dependent Analyses. Forest plot of the significant results of the period-dependent analyses. Multivariable logistic regression models were used to model the effect of each visit-dependent variable for protraction (P2-IM) (A) and fatigue at V3 (P2-fatigue) (B). A separate model was estimated for each predictor variable, with sex and age as additional covariables. The number of observations and AUC are provided in adjacent columns. HP indicates the medical history period prior to T_onset_, VEP the very early period from T_onset_ up to and including V1, and EP the early period from T_onset_ up to and including V2.

### Predictors for Chronic Fatigue (P2-fatigue)

Significant predictors of fatigue at V3 (P2-fatigue) of the *visit-*dependent analyses are shown in **Fig. 4B**. Fatigue was the most frequent IM feature at V3 (**Suppl. Fig. S3E**). It was, therefore, not unexpected to find overlaps between predictors for P2-fatigue and P2-IM. The best predictor for P2-fatigue regarding AUC was other airway symptoms which were significant risk factors after DAO 17, with subsequent OR increase over time. Moreover, moderate to severe GI symptoms and VCAp18 IgG detection in immunoblots were significant predictors with an increasing and decreasing OR over time, respectively (**Fig. 4B**). We did not find a significant *visit*-dependent effect of (very) early fatigue for P2-fatigue.

Significant predictors of fatigue at V3 (P2-fatigue) of the *period-*dependent analyses are shown in **Fig. 5B**. Early exanthema, early musculoskeletal symptoms, and ≥8 minor infections per year (ELVIS criterion) prior to IM significantly predicted P2-fatigue. Early IM_Clin_ complexity was the best predictor (AUC 0.74) of the *period-*dependent analyses. ROC analysis indicated that a cut-off of ≥12 IM features in the VEP (OR 2.43, 95%-CI: [1.07, 5.49]), and of ≥11 features in the EP (OR 3.39, 95%-CI: [1.41, 8.17]), as well as ≥9 IM_Clin_ features in both periods (VEP: OR 2.47, 95%-CI: [1.14, 5.33]; EP: OR 4.41, 95%-CI: [1.90, 10.21]) resulted in the greatest AUC for predicting fatigue at V3 (P2-fatigue), respectively. Early fatigue was not a significant predictor of P2-fatigue.

## DISCUSSION

Long-term sequelae of IM, such as chronic fatigue, manifest in a significant number of cases [19, 20] and hit young people in a very vulnerable stage of their lives, indicating a high medical need for research on risk factors, prevention, and therapeutic strategies [18, 19]. To identify biomarkers and predictors of acute and long-term IM complications we implemented the longitudinal IMMUC study and report here on the first results from a cohort of 200 pediatric and young adult patients.

In line with published data, 72.1% of the patients had fully recovered five to eight months after IM onset [14]. Remarkably, 27.9% of the study participants showed protracted disease for more than four months after IM onset, including 18.8% cases with chronic fatigue. Several clinical and laboratory parameters were identified that may predict protracted IM and thus identify patients at risk.

Of note, IM complexity was a significant predictor of long-term protraction. Within the VEP and EP, 50.5% and 94% study participants presented with maximum IM complexity. Ten or more very early IM_Clin_ features predicted protracted disease at V3 and nine or more predicted chronic fatigue. (Very) early IM_Lab_ features did not add any essential value to prediction accuracy.

Our findings are consistent with those reported by Cervia and colleagues, who identified the complexity of coronavirus disease 2019 (COVID-19) in combination with other clinical and laboratory characteristics as risk factors for post-acute COVID-19 syndromes [20]. It was hypothesized that a greater number of symptoms during the acute infection, including autonomic dysregulation, might reflect an initial immune (over)activation that results in prolonged impaired health [15, 21]. Our results are also in line with reports indicating an increased risk of ME/CFS in patients with more or more severe physical symptoms during the acute infection [13, 14].

Patients with distinct inborn errors of immunity (IEI) carry an increased susceptibility to EBV-associated HLH or lymphoproliferative disorders [3]. However, little is known about clinical IEI warning signs as possible predictors of protracted IM. In a previous study, 60% of ME/CFS patients reported a history of recurrent infections [22]. Our IMMUC data demonstrate that ≥1 major infection and ≥ 8 minor infections per year predicted any IM feature or fatigue at V3, respectively. These findings suggest that patients with an increased susceptibility to infection should be closely monitored and offered timely advice after IM diagnosis.

In addition, chronic fatigue was predicted by (very) early exanthema and facial edema, possibly indicative of IM-associated vascular inflammation [23-25], and was associated with early musculoskeletal symptoms and (very) early moderate to severe GI symptoms. Interestingly, GI symptoms prior to IM have been associated with the development of ME/CFS [15]. Taken together, these findings suggest that preexisting or early autonomic symptoms in the context of IM might predict post-viral disease.

Remarkably, the detection of EBV VCAp18 IgG considered a late marker of primary EBV infection [26], was identified as a significant predictor of any long-term protraction when detected within the first three weeks after IM onset. This might indicate that individuals with a long EBV incubation period prior to IM may face an increased risk of protracted disease, and early immunoblot analysis might help in prediction. Early detection of EBV VCAp18 IgG has been associated with a lower IM severity score by others but was not investigated as a predictor of protracted disease [27]. Moreover, EBV VCAp18 IgM-mediated complement deposition and EBV VCAp18 IgG-dependent cellular phagocytosis were positively associated with IM severity [28], and complement activation was discussed in ME/CFS-associated immune dysregulation [22].

### Limitations

First, the results of the IMMUC scores might have been prone to interviewer bias. Second, most patients were recruited when initial IM symptoms had declined. Therefore, laboratory parameters at V1 may reflect a subacute stage of IM and remembered IM symptoms of the VEP might be influenced by recall bias. Third, the time period between V1 and follow-up visits varied to match best with school schedules and family plans.

## Conclusion

In this IMMUC study, we implemented a novel scoring system to evaluate IM severity, complexity, and protraction and identified several candidate predictors of chronic postviral disease that are easily assessed during routine diagnostics. This score may help identifying patients at risk and facilitate prompt medical provisioning. In addition, this score forms the basis for the analysis of diverse immunological parameters investigated in this cohort that might further improve prediction for long-term sequelae in young people with IM.

## Data Availability

All data produced in the present study is not openly available.

## Acknowledgments

The authors would like to thank the IMMUC Study Group team members F. Fischer, M. Burggraf, P. Wallraven, R. Weggel, K. Bartl, L. Werny, K. Rautter, T. Hofberger, Y. Müller, H. Zietemann, E. Planatscher, J. Nückel, K. Michel, S. Strunz., J. Mücke, K. Mittelstraß, M. Bach, F. Martin, L. Kramer, C. Richter, A. Yardim, and L. Beckert for their great study assistance. Furthermore we would like to thank patients and their families for participating in the study, as well as all participating physicians for announcing the study and transferring interested patients. Without their tremendous support this work would have not been possible.

## Author contributions

U.B. acquired funding, supervised and coordinated the study together with K.G.. U.B., M.B., J.G., A.H., S.S, N.K., T.B., J.M, D.H., P.L., S.E-S., H-J.D., S.D., F.H., C.F., T.S., M-M.S., A.B., and K.G. designed the study. M.B., J.G., L.S-H., R.P., F.H., C.F., M-M.S., A.B., K.G., and U.B. recruited and visited the patients and acquired the data. M.B., J.G., J.L.L., J.W., A.H., E.N., F.H., M-M.S., A.B., A.N., L.M., K.G., and U.B. analysed and interpreted the data. M.B., J.G., L.M., J.L.L., A.H., A.N., K.G., and U.B. wrote the first version of the manuscript. All authors read, edited, and commented on the draft. All authors approved the final version of the manuscript to be published and agreed to be accountable for all aspects of the work in ensuring that questions related to the accuracy or integrity of any part of the work are appropriately investigated and resolved.

## Availability of data and materials

M.B., J.G., L.M., J.L.L., A.H., J.W., K.G., and U.B. have full access to all data and take responsibility for their integrity and accuracy.

## Ethical considerations

The IMMUC study was approved by the Ethics Committee of the School of Medicine and Health of the Technical University of Munich (reference number: 112/14).

## SUPPLEMENTARY METHODS

### Study Data

Baseline patient characteristics included age, sex, body mass index (BMI), body height, serum titers (anti-diphtheria and anti-tetanus toxoid IgG, total IgA, IgM, IgG, IgE, vitamin D), and data from the general medical history of patients and families (**Suppl. Tab. 1**). The patient’s medical history addressed clinical signs of pathological susceptibility to infections (ELVIS) [17] and immune dysregulation (GARFIELD) [17], failure to thrive, developmental delay, vaccination status, infectious diseases (influenza, herpes labialis, varicella, measles), allergies, asthma, cancer, transplantation, transfusions, exposure to farm animals, as well as specific events within 6 months before T_onset_ (fever, use of antibiotics/analgesics/ antiphlogistics, traveling to certain regions). The family’s medical history addressed parent consanguinity, the number of siblings in the same household, as well as a history of IM, known inborn errors of immunity (IEI), cancer, neurodermatitis, autoimmune disease, allergies, and/or asthma (**Suppl. Mat. M1**). IM_Clin_ features included fever, fatigue, lymphadenopathy, tonsillopharyngitis, splenomegaly, facial edema, exanthema, other airway symptoms, other gastrointestinal (GI) symptoms, urogenital symptoms, bleeding and/or treatment for bleeding or anemia, neurological symptoms, musculoskeletal symptoms, hospitalization, and treatment for HLH (**Suppl. Mat. M2**); they were evaluated retrospectively and prospectively from T_onset_ until V3. IM_Lab_ features included lymphocytosis, neutropenia, anemia, thrombocytopenia, hyperferritinemia, hepatitis, nephropathy, and CRP elevation defined by age-adapted cut-off levels of blood count, transaminases (ASAT, ALAT), CRP, ferritin, creatinine, and (cystatin C if creatinine levels were elevated) in serum. IM_Vir_ features included EBV DNA in plasma (EBV_Plasma_), cell fraction (EBV_cells_), and oral washes (EBV_oral_) evaluated by PCR (TaqManTM 7500, Thermo Fisher Scientific, Waltham, USA), as well as anti-EBV VCA IgM, anti-EBV VCA IgG, and anti-EBNA-1 IgG determined in serum by CMIA [ARCHITECT i1000SR^TM^, Abbot Laboratories, Chicago, USA]), and serum IgG against EAp54, EAp138, VCAp18, VCAp23, BZLF1 or EBNA-1 detected by immunoblot (ProfiBlotTM, Tecan Group Ltd., Männedorf, Switzerland).

### Sample Size Calculation

With 20 expected P2 cases and a required minimum number of 5-10 observations in the less frequent outcome class, we could consistently estimate multivariable logistic regression models with 2-4 parameters [31, 32]. An unpaired two-sided t-test with a 5% significance level has 80% power to detect an effect size of 0.7 when the sample sizes in two groups are N=18 (P2) and N=180 (P0-1) in a total sample size of N=198, as expected (nQuery Advisor 7.0).

### SUPPLEMENTARY TABLES

**Supplementary Table S1A.** Baseline Patient Characteristics – Sex and Age Distribution at Recruitment.

| Sex | Number/Total (%) |
| --- | --- |
| Male | 91/200 (45.5%) |
| Female | 109/200 (54.5%) |
| Age Groups | Number/Total (%) |
| Children 0-11 years | 92/200 (46.0%) |
| Adolescents 12-17 years | 84/200 (42.0%) |
| Adults 18-39 years | 24/200 (12.0%) |

**Supplementary Table S1B.** Baseline Patient Characteristics – Body Measures and Serum Titers.

| Characteristics | Number/Total (%) |  |  |
| --- | --- | --- | --- |
|  | Below Normal Range | Within Normal Range | Above Normal Range |
| Body height [1] | 4/198 (2.0%) | 182/198 (91.9%) | 12/198 (6.1%) |
| Body mass index (BMI) [1] | 9/200 (4.5%) | 188/200 (94.0%) | 3/200 (1.5%) |
| Vitamin D [2] | 68/200 (34.0%) | 132/200 (66.0%) | 0/200 (0.0%) |
| Total IgE [2] | n/a | 111/200 (55.5%) | 89/200 (44.5%) |
| Total IgA [2] | 0/200 (0.0%) | 156/200 (78.0%) | 44/200 (22.0%) |
| Total IgM [2] | 3/200 (1.5%) | 129/200 (64.5%) | 68/200 (34.0%) |
| Total IgG [2] | 3/200 (1.5%) | 162/200 (81.0%) | 35/200 (17.5%) |
| Anti-Diphtheria toxoid IgG [3] | 0/200 (0.0%) | 200/200 (100.0%) | n/a |
| Anti-Tetanus toxoid IgG [3] | 8/200 (4%) | 192/200 (96.0%) | n/a |
| Magnesium [2] | 0/170 (0.0%) | 143/170 (84.1%) | 27/170 (15.9%) |

n/a: not applicable
[1] Age-adapted normal range of body measures (3<sup>th</sup> – 97<sup>th</sup> percentile) [33-35]
[2] Normal range vitamin D serum, total immunoglobulins (Ig) (5<sup>th</sup> – 95<sup>th</sup> percentile) and magnesium serum [36]
[3] Normal range of vaccination titers according to the information of the standing committee on vaccination (STIKO) at the Robert Koch Institute (RKI), a department of the German Federal Health Agency [37], accessed on February 6<sup>th</sup>, 2024.

**Supplementary Table S1C.** Baseline Patient Characteristics – General Medical History.

| Vaccination Status | Number/Total (%) |
| --- | --- |
| Complete vaccination status [4] | 177/199 (88.9%) |
| Incomplete vaccination status [4] | 22/199 (11.1%) |
| Specific Infectious Diseases | Number/Total (%) |
| Herpes labialis | 9/199 (4.5%) |
| Influenza | 12/199 (6.0%) |
| Measles | 5/199 (2.5%) |
| Varizella (chickenpox) | 74/199 (37.2%) |
| Medical Interventions | Number/Total (%) |
| Transfusion | 3/200 (1.5%) |
| Transplantation | 1/200 (0.5%) |
| Others | Number/Total (%) |
| Allergies | 58/200 (29.0%) |
| Asthma | 16/200 (8.0%) |
| Developmental delay | 1/200 (0.5%) |
| Type 1 diabetes mellitus | 1/200 (0.5%) |
| Henoch-Schoenlein purpura | 1/200 (0.5%) |
| Immune thrombocytopenic purpura | 1/200 (0.5%) |
| Known failure to thrive | 3/200 (1.5%) |
| Lichen sclerosis | 1/200 (0.5%) |
| Malignant tumor | 1/200 (0.5%) |
| Neurodermatitis | 15/200 (7.5%) |
| Rheumatoid arthritis | 1/200 (0.5%) |

[4] Vaccination status according to the recommendations of the standing committee on vaccination (STIKO) at the Robert Koch Institute (RKI), a department of the German Federal Health Agency [38], accessed at the beginning of patient recruitment on February 11<sup>th</sup>, 2016.

**Supplementary Table S1D.**
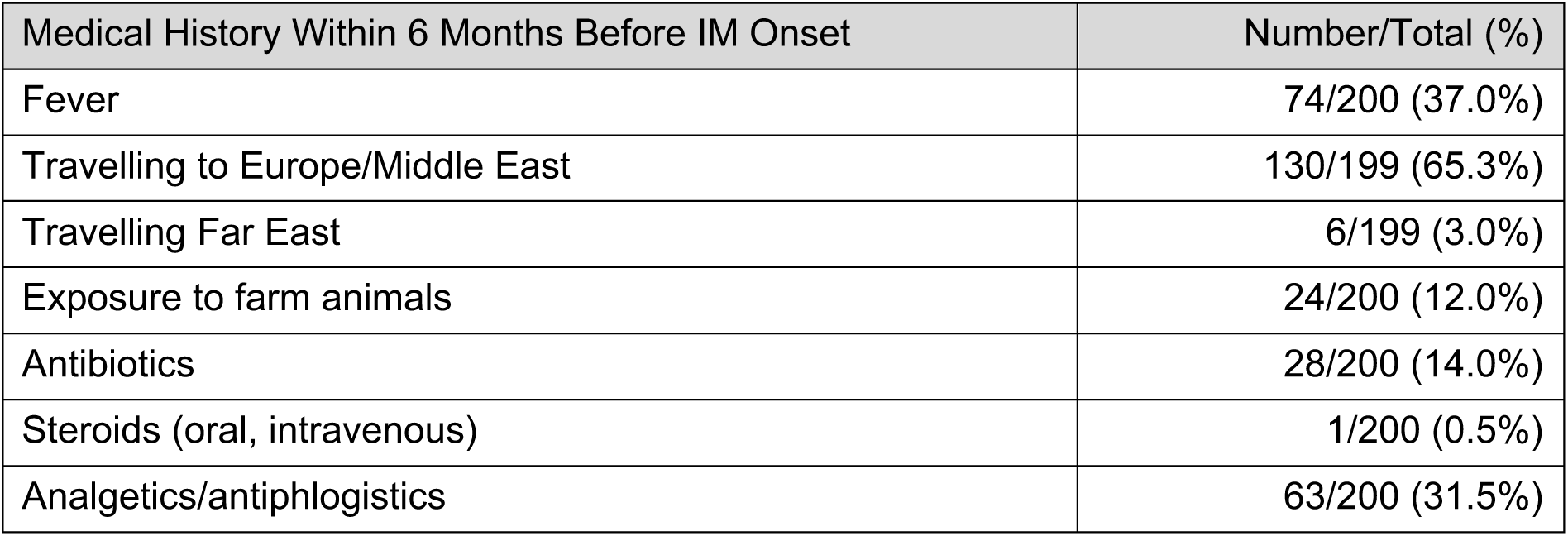
Baseline Patient Characteristics – Recent Medical History.

| Medical History Within 6 Months Before IM Onset | Number/Total (%) |
| --- | --- |
| Fever | 74/200 (37.0%) |
| Travelling to Europe/Middle East | 130/199 (65.3%) |
| Travelling Far East | 6/199 (3.0%) |
| Exposure to farm animals | 24/200 (12.0%) |
| Antibiotics | 28/200 (14.0%) |
| Steroids (oral, intravenous) | 1/200 (0.5%) |
| Analgetics/antiphlogistics | 63/200 (31.5%) |

**Supplementary Table S1E.** Baseline Patient Characteristics – Signs of Primary Immunodeficiency.

| Signs of Primary Immunodeficiency | Number/Total (%) |
| --- | --- |
| Known primary immunodeficiency | 1/200 (0.5%) |
| ELVIS – at least one criterion [5] | 77/200 (38.5%) |
| ELVIS – one criterion | 60/200 (30.0%) |
| ELVIS – two criteria | 13/200 (6.5%) |
| ELVIS – three criteria | 3/200 (1.5%) |
| ELVIS – four criteria | 1/200 (0.5%) |
| ELVIS – unusual pathogen | 8/200 (4.0%) |
| ELVIS – multiple localization | 4/199 (2.0%) |
| ELVIS – relapsing course | 29/200 (14.5%) |
| ELVIS – unusual intensity | 33/200 (16.5%) |
| ELVIS – increased sum of minor infections | 25/200 (12.5%) |
| GARFIELD – at least one criterion [5] | 37/200 (18.5%) |
| GARFIELD – one criterion | 35/200 (17.5%) |
| GARFIELD – two criteria | 2/200 (1.0%) |
| GARFIELD – granuloma | 0/200 (0.0%) |
| GARFIELD – autoimmunity | 5/200 (2.5%) |
| GARFIELD – recurring fever | 7/200 (3.5%) |
| GARFIELD – eczema, including neurodermatitis | 23/200 (11.5%) |
| GARFIELD – lymphoproliferation | 3/200 (1.5%) |
| GARFIELD – chronic inflammatory bowel disease | 1/200 (0.5%) |
| ELVIS or GARFIELD – at least one criterion [5] | 95/200 (47.5%) |
| ELVIS and GARFIELD – at least one criterion of both [5] | 19/200 (9.5%) |

**Supplementary Table S1F.** Baseline Patient Characteristics – General Medical History of Family.

| Characteristics | Number/Total (%) |
| --- | --- |
| Consanguineous parents | 3/200 (1.5%) |
| At least 1 sibling in the same household | 158/200 (79.4%) |
| 1 sibling | 108/200 (54.0%) |
| 2 siblings | 36/200 (18.0%) |
| ≥ 3 siblings | 14/200 (7.0%) |
| Allergies | 125/199 (62.8%) |
| Allergic asthma | 15/200 (7.5%) |
| Infectious mononucleosis | 71/199 (35.7%) |
| 1 family member | 59/199 (29.6%) |
| ≥ 2 family members | 12/199 (6.0%) |
| Neurodermatitis | 29/199 (14.6%) |
| Classical autoimmune diseases | 41/200 (20.5%) |
| Cancer | 126/199 (63.3%) |
| Primary immunodeficiencies | 1/199 (0.5%) |

**Suppl. Table S2.** Severity of IM features at Study Visits and Study Periods.

| IM Feature | Early Period (EP) | Very Early Period (VEP) | Visit 1 (V1) | Visit 2 (V2) | Visit 3 (V3) |
| --- | --- | --- | --- | --- | --- |
|  | Number/Total (%) | Number/Total (%) | Number/Total (%) | Number/Total (%) | Number/Total (%) |
| **IM<sub>Clin</sub> Features** |  |  |  |  |  |
| **Fever** |  |  |  |  |  |
| S0 | 55/200 (27.5%) | 59/199 (29.6%) | 164/196 (83.7%) | 190/191 (99.5%) | 178/179 (99.4%) |
| S1 | 55/200 (27.5%) | 55/199 (27.6%) | 23/196 (11.7%) | 0/191 (0.0%) | 1/179 (0.6%) |
| S2 | 90/200 (45.0%) | 85/199 (42.7%) | 9/196 (4.6%) | 1/191 (0.5%) | 0/179 (0.0%) |
| **Fatigue** |  |  |  |  |  |
| S0 | 8/200 (4.00%) | 14/200 (7.0%) | 34/200 (17.0%) | 102/193 (52.8%) | 147/181 (81.2%) |
| S1 | 143/200 (71.5%) | 142/200 (71.0%) | 154/200 (77.0%) | 91/193 (47.2%) | 28/181 (15.5%) |
| S2 | 42/200 (21.0%) | 38/200 (19.0%) | 12/200 (6.0%) | 0/193(0.0%) | 6/181 (3.3%) |
| S3 | 7/200 (3.5%) | 6/200 (3.0%) | 0/200 (0.0%) | 0/193(0.0%) | 0/181 (0.0%) |
| **Lymphadenopathy** |  |  |  |  |  |
| S0 | 17/200 (8.5%) | 20/200 (10.0%) | 35/200 (17.5%) | 126/192 (66.6%) | 158/173 (91.3%) |
| S1 | 135/200 (67.5%) | 134/200 (67.0%) | 143/200 (71.5%) | 66/192 (34.4%) | 15/173 (8.7%) |
| S2 | 44/200 (22.0%) | 43/200 (21.5%) | 21/200 (10.5%) | 0/192 (0.0%) | 0/173 (0.0%) |
| S3 | 2/200 (1.0%) | 2/200 (1.0%) | 1/200 (0.5%) | 0/192 (0.0%) | 0/173 (0.0%) |
| S4 | 2/200 (1.0%) | 1/200 (0.5%) | 0/200 (0.0%) | 0/192 (0.0%) | 0/173 (0.0%) |
| **Tonsillopharyngitis** |  |  |  |  |  |
| S0 | 17/200 (8.5%) | 20/200 (10.0%) | 46/200 (23.0%) | 166/192 (86.5%) | 159/172 (92.4%) |
| S1 | 96/200 (48.0%) | 94/200 (47.0%) | 105/200 (52.5%) | 25/192 (13.0%) | 13/172 (7.6%) |
| S2 | 63/200 (31.5%) | 63/200 (31.5%) | 39/200 (19.5%) | 1/192 (0.5%) | 0/172 (0.0%) |
| S3 | 21/200 (10.5%) | 20/200 (10.0%) | 7/200 (3.5%) | 0/192 (0.0%) | 0/172 (0.0%) |
| S4 | 3/200 (1.5%) | 3/200 (1.5%) | 3/200 (1.5%) | 0/192(0.0%) | 0/172 (0.0%) |
| Splenomegaly |  |  |  |  |  |
| S0 | 48/195 (24.6%) | 48/189 (25.4%) | 51/186 (27.4%) | 111/183 (60.7%) | 26/37 (70.3%) |
| S1 | 90/195 (46.2%) | 89/189 (47.1%) | 84/186 (45.2%) | 60/183(32.8%) | 11/37 (29.7%) |
| S2 | 56/195 (28.7%) | 51/189 (27.0%) | 50/186 (26.9%) | 12/183 (6.6%) | 0/37 (0.0%) |
| S4 | 1/195 (0.5%) | 1/189 (0.5%) | 1/186 (0.5%) | 0/183 (0.0%) | 0/37 (0.0%) |
| Facial Edema |  |  |  |  |  |
| S0 | 113/200 (56.5%) | 114/200 (57.0%) | 150/200 (75.0%) | 191/194 (98.5%) | 181/183 (98.9%) |
| S1 | 60/200 (30.0%) | 61/200 (30.5%) | 41/200 (20.5%) | 3/194 (1.5%) | 2/183 (1.1%) |
| S2 | 22/200 (11.0%) | 21/200 (10.5%) | 7/200 (3.5%) | 0/194 (0.0%) | 0/183 (0.0%) |
| S3 | 5/200 (2.5%) | 4/200 (2.0%) | 2/200 (1.0%) | 0/194 (0.0%) | 0/183 (0.0%) |
| Exanthema |  |  |  |  |  |
| S0 | 145/200 (72.5%) | 154/198 (77.8%) | 171/197 (86.8%) | 181/190 (95.3%) | 179/180 (99.4%) |
| S1 | 28/200 (14.0%) | 21/198 (10.6%) | 9/197 (4.6%) | 6/190 (3.2%) | 1/180 (0.6%) |
| S2 | 21/200 (10.5%) | 17/198 (8.6%) | 13/197 (6.6%) | 3/190 (1.6%) | 0/180 (0.0%) |
| S3 | 6/200 (3.0%) | 6/198 (3.0%) | 4/197 (2.0%) | 0/190 (0.0%) | 0/180 (0.0%) |
| S4 | 0/200 (0.0%) | 0/198 (0.0%) | 0/197 (0.0%) | 0/190 (0.0%) | 0/180 (0.0%) |
| Bleeding and/or Treatment for Bleeding or Anemia |  |  |  |  |  |
| S0 | 189/200 (94.5%) | 190/200 (95.0%) | 196/198 (99.0%) | 192/193 (99.5%) | 181/181 (100.0%) |
| S1 | 11/200 (5.5%) | 10/200 (5.0%) | 2/198 (1.0%) | 1/193 (0.5%) | 0/181 (0.0%) |
| S2 | 0/200 (0.0%) | 0/200 (0.0%) | 0/198 (0.0%) | 0/193 (0.0%) | 0/181 (0.0%) |
| S3 | 0/200 (0.0%) | 0/200 (0.0%) | 0/198 (0.0%) | 0/193 (0.0%) | 0/181 (0.0%) |
| S4 | 0/200 (0.0%) | 0/200 (0.0%) | 0/198 (0.0%) | 0/193 (0.0%) | 0/181 (0.0%) |
| Other Airway Symptoms |  |  |  |  |  |
| S0 | 41/199 (20.6%) | 43/196 (21.9%) | 79/194 (40.7%) | 162/191 (84.8%) | 151/162 (93.2%) |
| S1 | 141/199 (70.9%) | 137/196 (69.9%) | 108/194 (55.7%) | 28/191 (14.7%) | 11/162 (6.8%) |
| S2 | 7/199 (3.5%) | 7/196 (3.6%) | 3/194 (1.5%) | 1/191 (0.5%) | 0/162 (0.0%) |
| S3 | 8/199 (4.0%) | 8/196 (4.1%) | 4/194 (2.1%) | 0/191 (0.0%) | 0/162 (0.0%) |
| S4 | 2/199 (1.0%) | 1/196 (0.5%) | 0/194 (0.0%) | 0/191 (0.0%) | 0/162 (0.0%) |
| Other Gastrointestinal Symptoms |  |  |  |  |  |
| S0 | 57/200 (28.5%) | 66/199 (33.2%) | 132/198 (66.7%) | 171/191 (89.5%) | 170/177 (96.0%) |
| S1 | 121/200 (60.5%) | 116/199 (58.3%) | 57/198 (28.8%) | 18/191 (9.4%) | 7/177 (4.0%) |
| S2 | 10/200 (5.0%) | 8/199 (4.0%) | 3/198 (1.5%) | 1/191 (0.5%) | 0/177 (0.0%) |
| S3 | 12/200 (6.0%) | 9/199 (4.5%) | 6/198 (3.0%) | 1/191 (0.5%) | 0/177 (0.0%) |
| S4 | 0/200 (0.0%) | 0/199 (0.0%) | 0/198 (0.0%) | 0/191 (0.0%) | 0/177 (0.0%) |
| Urogenital Symptoms |  |  |  |  |  |
| S0 | 187/199 (94.0%) | 185/197 (93.9%) | 192/197 (97.5%) | 191/193 (99.0%) | 178/180 (98.9%) |
| S1 | 10/199 (5.0%) | 10/197 (5.1%) | 4/197 (2.0%) | 2/193 (1.0%) | 2/180 (1.1%) |
| S2 | 2/199 (1.0%) | 2/197 (1.0%) | 1/197 (0.5%) | 0/193 (0.0%) | 0/180 (0.0%) |
| S3 | 0/199 (0.0%) | 0/197 (0.0%) | 0/197 (0.0%) | 0/193 (0.0%) | 0/180 (0.0%) |
| S4 | 0/199 (0.0%) | 0/197 (0.0%) | 0/197 (0.0%) | 0/193 (0.0%) | 0/180 (0.0%) |
| Neurological Symptoms |  |  |  |  |  |
| S0 | 56/199 (28.1%) | 62/196 (31.6%) | 135/196 (68.9%) | 172/191 (90.0%) | 166/179 (92.7%) |
| S1 | 138/199 (69.3%) | 129/196 (65.8%) | 58/196 (29.6%) | 18/191 (9.4%) | 13/179 (7.3%) |
| S2 | 2/199 (1.0%) | 2/196 (1.0%) | 2/196 (1.0%) | 0/191 (0.0%) | 0/179 (0.0%) |
| S3 | 1/199 (0.5%) | 1/196 (0.5%) | 0/196 (0.0%) | 0/191 (0.0%) | 0/179 (0.0%) |
| S4 | 2/199 (1.0%) | 2/196 (1.0%) | 1/196 (0.5%) | 1/191 (0.5%) | 0/179 (0.0%) |
| Musculoskeletal Symptoms |  |  |  |  |  |
| S0 | 103/199 (51.8%) | 123/197 (62.4%) | 168/197 (85.3%) | 180/192 (93.8%) | 174/178 (97.8%) |
| S1 | 83/199 (41.7%) | 63/197 (32.0%) | 24/197 (12.2%) | 12/192 (6.3%) | 4/178 (2.2%) |
| S2 | 11/199 (5.5%) | 9/197 (4.6%) | 5/197 (2.5%) | 0/192 (0.0%) | 1/178 (0.6%) |
| S3 | 2/199 (1.0%) | 2/197 (1.0%) | 0/197 (0.0%) | 0/192 (0.0%) | 0/178 (0.0%) |
| Imminent or Treatment for Hemaphagocytic Lymphohistiocytosis (HLH) |  |  |  |  |  |
| S0 | 199/200 (99.5%) | 199/200 (99.5%) | 199/200 (99.5%) | 193/194 (99.5%) | 182/183 (99.5%) |
| S3 | 0/200 (0.0%) | 0/200 (0.0%) | 0/200 (0.0%) | 1/194 (0.5%) | 1/183 (0.5%) |
| S4 | 1/200 (0.5%) | 1/200 (0.5%) | 1/200 (0.5%) | 0/194 (0.0%) | 0/183 (0.0%) |
| Hospitalization |  |  |  |  |  |
| S0 | 111/200 (55.5%) | 115/200 (57.5%) | 134/200 (67.0%) | 194/194 (100.0%) | 181/182 (99.5%) |
| S2 | 81/200 (40.5%) | 81/200 (40.5%) | 62/200 (31.0%) | 0/194 (0.0%) | 0/182 (0.0%) |
| S3 | 7/200 (3.5%) | 3/200 (1.5%) | 4/200 (2.0%) | 0/194 (0.0%) | 1/182 (0.5%) |
| S4 | 1/200 (0.5%) | 1/200 (0.5%) | 0/200 (0.0%) | 0/194 (0.0%) | 0/182 (0.0%) |
| IM<sub>Lab</sub> Features |  |  |  |  |  |
| Lymphocytosis |  |  |  |  |  |
| S0 | 155/200 (77.5%) | 164/200 (82.0%) | 178/195 (91.3%) | 183/185 (98.9%) | 175/175 (100.0%) |
| S1 | 45/200 (22.5%) | 36/200 (18.0%) | 17/195 (8.7%) | 2/185 (1.1%) | 0/175 (0.0%) |
| Neutropenia |  |  |  |  |  |
| S0 | 116/198 (58.6%) | 151/197 (76.6%) | 159/192 (82.8%) | 157/183 (85.8%) | 170/173 (98.3%) |
| S1 | 54/198 (27.3%) | 35/197 (17.8%) | 28/192 (14.6%) | 22/183 (12.0%) | 2/173 (1.2%) |
| S2 | 27/198 (13.6%) | 10/197 (5.1%) | 5/192 (2.6%) | 4/183 (2.2%) | 1/173 (0.6%) |
| S3 | 1/198 (0.5%) | 1/197 (0.5%) | 0/192 (0.0%) | 0/183 (0.0%) | 0/173 (0.0%) |
| Anemia |  |  |  |  |  |
| S0 | 190/198 (96.0%) | 190/197 (96.4%) | 187/192 (97.4%) | 182/184 (98.9%) | 173/174 (99.4%) |
| S1 | 3/198 (1.5%) | 3/197 (1.5%) | 3/192 (1.6%) | 1/184 (0.5%) | 1/174 (0.6%) |
| S2 | 5/198 (2.5%) | 4/197 (2.0%) | 2/192 (1.0%) | 1/184 (0.5%) | 0/174 (0.0%) |
| S3 | 0/198 (0.0%) | 0/197 (0.0%) | 0/192 (0.0%) | 0/184 (0.0%) | 0/174 (0.0%) |
| S4 | 0/198 (0.0%) | 0/197 (0.0%) | 0/192 (0.0%) | 0/184 (0.0%) | 0/174 (0.0%) |
| Thrombocytopenia |  |  |  |  |  |
| S0 | 148/199 (74.4%) | 151/199 (75.9%) | 176/194 (90.7%) | 184/186 (98.9%) | 173/175 (98.9%) |
| S1 | 50/199 (25.1%) | 47/199 (23.6%) | 17/194 (8.8%) | 1/186 (0.5%) | 2/175 (1.1%) |
| S2 | 0/199 (0.0%) | 0/199 (0.0%) | 0/194 (0.0%) | 0/186 (0.0%) | 0/175 (0.0%) |
| S3 | 0/199 (0.0%) | 0/199 (0.0%) | 0/194 (0.0%) | 0/186 (0.0%) | 0/175 (0.0%) |
| S4 | 1/199 (0.5%) | 1/199 (0.5%) | 1/194 (0.5%) | 1/186 (0.5%) | 0/175 (0.0%) |
| Hyperferritinemia |  |  |  |  |  |
| S0 | 124/200 (62.0%) | 122/192 (63.5%) | 129/195 (66.2%) | 180/186 (96.8%) | 175/176 (99.4%) |
| S1 | 56/200 (28.0%) | 51/192 (26.6%) | 50/195 (25.6%) | 5/186 (2.7%) | 0/176 (0.0%) |
| S2 | 15/200 (7.5%) | 14/192 (7.3%) | 14/195 (7.2%) | 1/186 (0.5%) | 1/176 (0.6%) |
| S3 | 4/200 (2.0%) | 4/192 (2.1%) | 1/195 (0.5%) | 0/186 (0.0%) | 0/176 (0.0%) |
| S4 | 1/200 (0.5%) | 1/192 (0.5%) | 1/195 (0.5%) | 0/186 (0.0%) | 0/176 (0.0%) |
| Hepatitis or Cholangitis |  |  |  |  |  |
| S0 | 70/200 (35.0%) | 74/200 (37.0%) | 86/196 (43.9%) | 155/186 (83.3%) | 174/174 (100.0%) |
| S1 | 105/200 (52.5%) | 101/200 (50.5%) | 97/196 (49.5%) | 31/186 (16.7%) | 0/174 (0.0%) |
| S2 | 19/200 (9.5%) | 19/200 (9.5%) | 10/196 (5.1%) | 0/186 (0.0%) | 0/174 (0.0%) |
| S3 | 6/200 (3.0%) | 6/200 (3.0%) | 3/196 (1.5%) | 0/186 (0.0%) | 0/174 (0.0%) |
| S4 | 0/200 (0.0%) | 0/200 (0.0%) | 0/196 (0.0%) | 0/186 (0.0%) | 0/174 (0.0%) |
| Nephropathy |  |  |  |  |  |
| S0 | 197/199 (99.0%) | 194/195 (99.5%) | 193/194 (99.5%) | 186/186 (100.0%) | 173/173 (100.0%) |
| S1 | 1/199 (0.5%) | 0/195 (0.0%) | 0/194 (0.0%) | 0/186 (0.0%) | 0/173 (0.0%) |
| S2 | 1/199 (0.5%) | 1/195 (0.5%) | 1/194 (0.5%) | 0/186 (0.0%) | 0/173 (0.0%) |
| S3 | 0/199 (0.0%) | 0/195 (0.0%) | 0/194 (0.0%) | 0/186 (0.0%) | 0/173 (0.0%) |
| Elevation of C-reactive Protein |  |  |  |  |  |
| S0 | 83/200 (41.5%) | 86/200 (43.0%) | 123/196 (62.8%) | 180/187 (96.3%) | 175/175 (100.0%) |
| S1 | 106/200 (53.0%) | 105/200 (52.5%) | 72/196 (36.7%) | 7/187 (3.7%) | 0/175 (0.0%) |
| S2 | 11/200 (5.5%) | 9/200 (4.5%) | 1/196 (0.5%) | 0/187 (0.0%) | 0/175 (0.0%) |
| S3 | 0/200 (0.0%) | 0/200 (0.0%) | 0/196 (0.0%) | 0/187 (0.0%) | 0/175 (0.0%) |

### SUPPLEMENTARY FIGURE LEGENDS

**Supplementary Figure S1.**
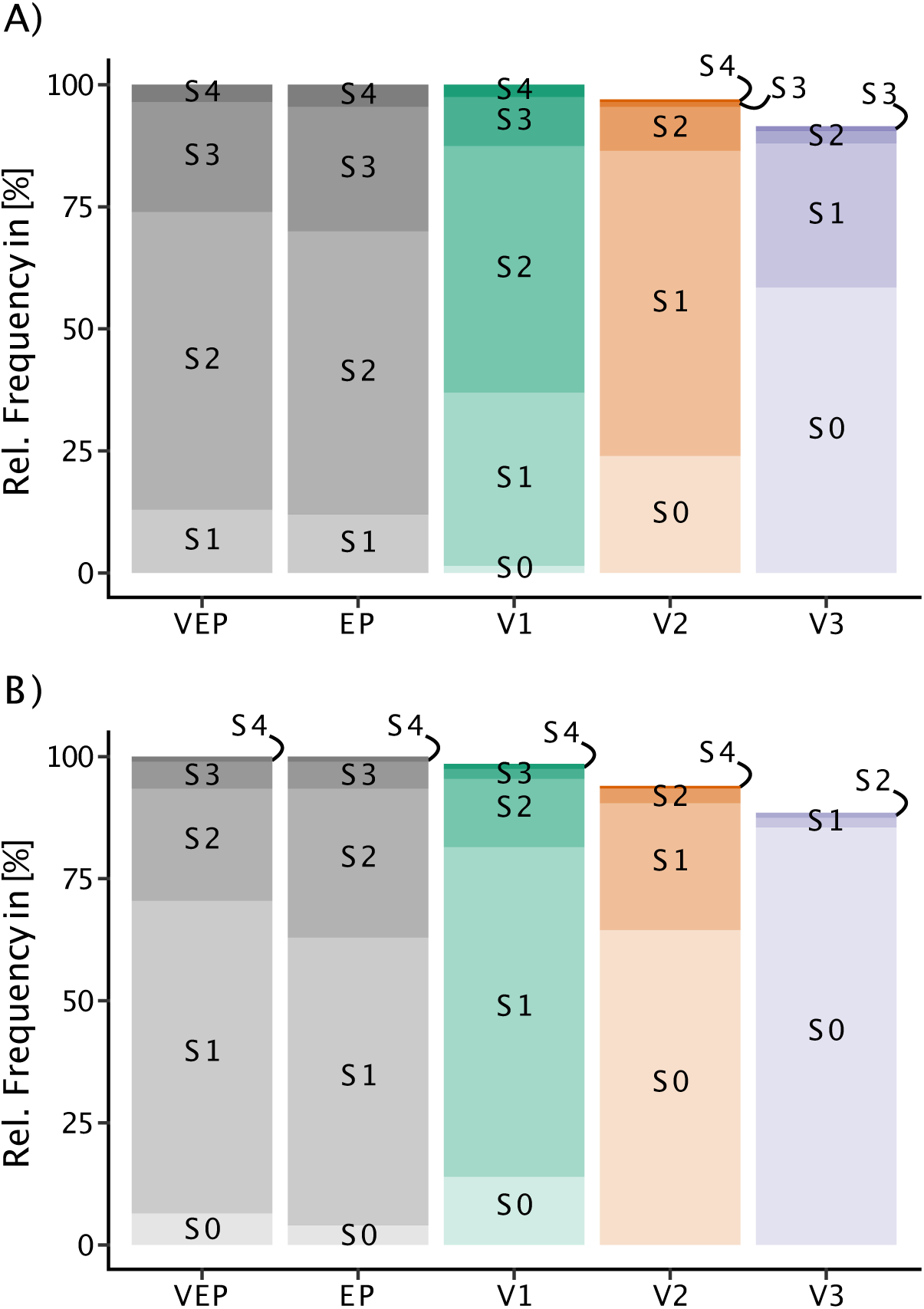
Clinical and Laboratory IMMUC Severity Scores. The relative frequency of the severity of clinical (IM_Clin_) (A) and laboratory symptoms (IM_Lab_) (B) for the very early period (VEP, grey), the early period (EP, grey), and the visits 1 – 3 (colored) are shown.

**Supplementary Figure S2.**
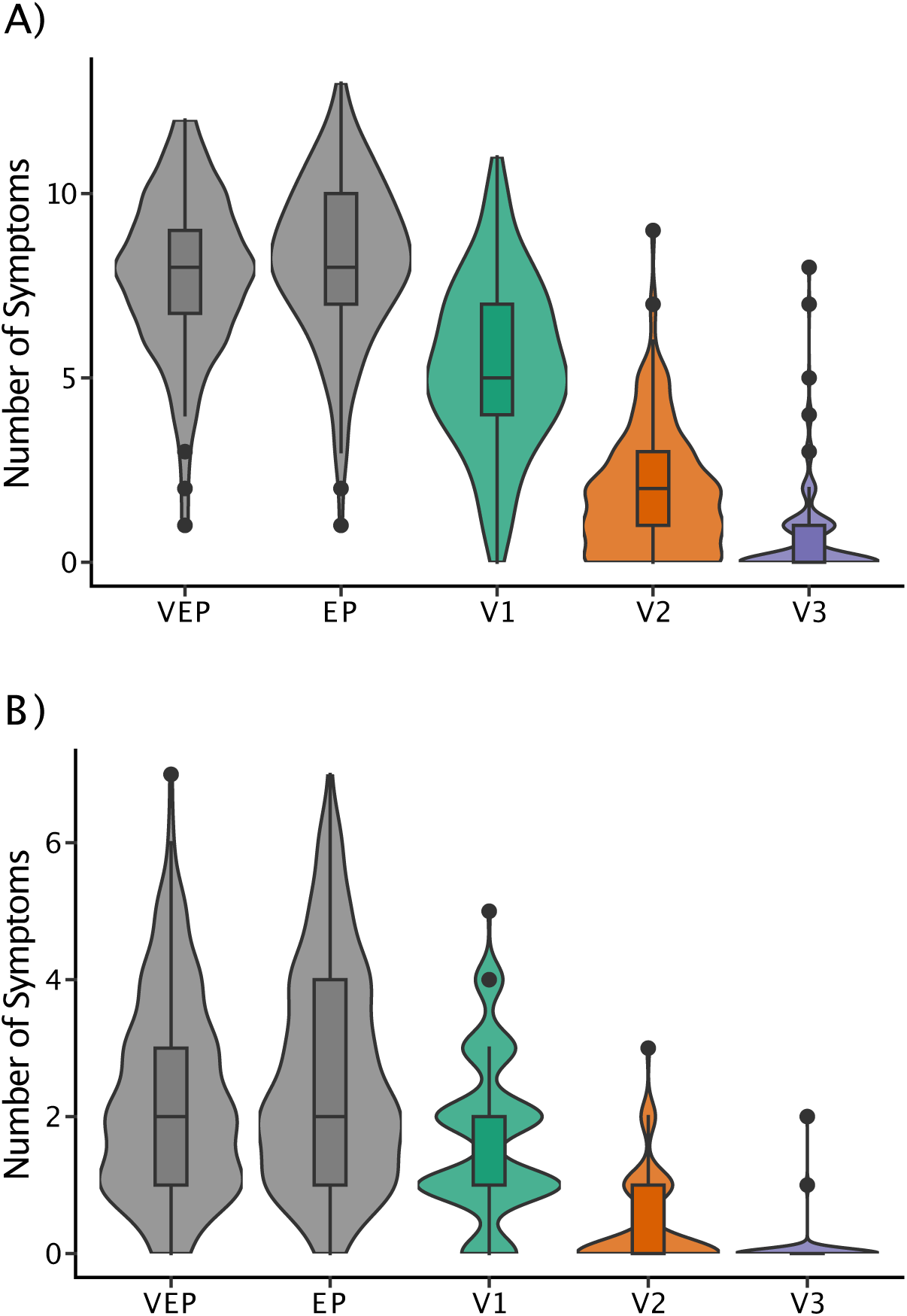
Clinical and Laboratory IMMUC Complexity Scores. The complexity of clinical (IM_Clin_) (A) and laboratory symptoms (IM_Lab_) (B) during the very early period (VEP, grey), the early period (EP, grey), and the visits 1 – 3 (colored) are shown.

**Supplementary Figure S3.**
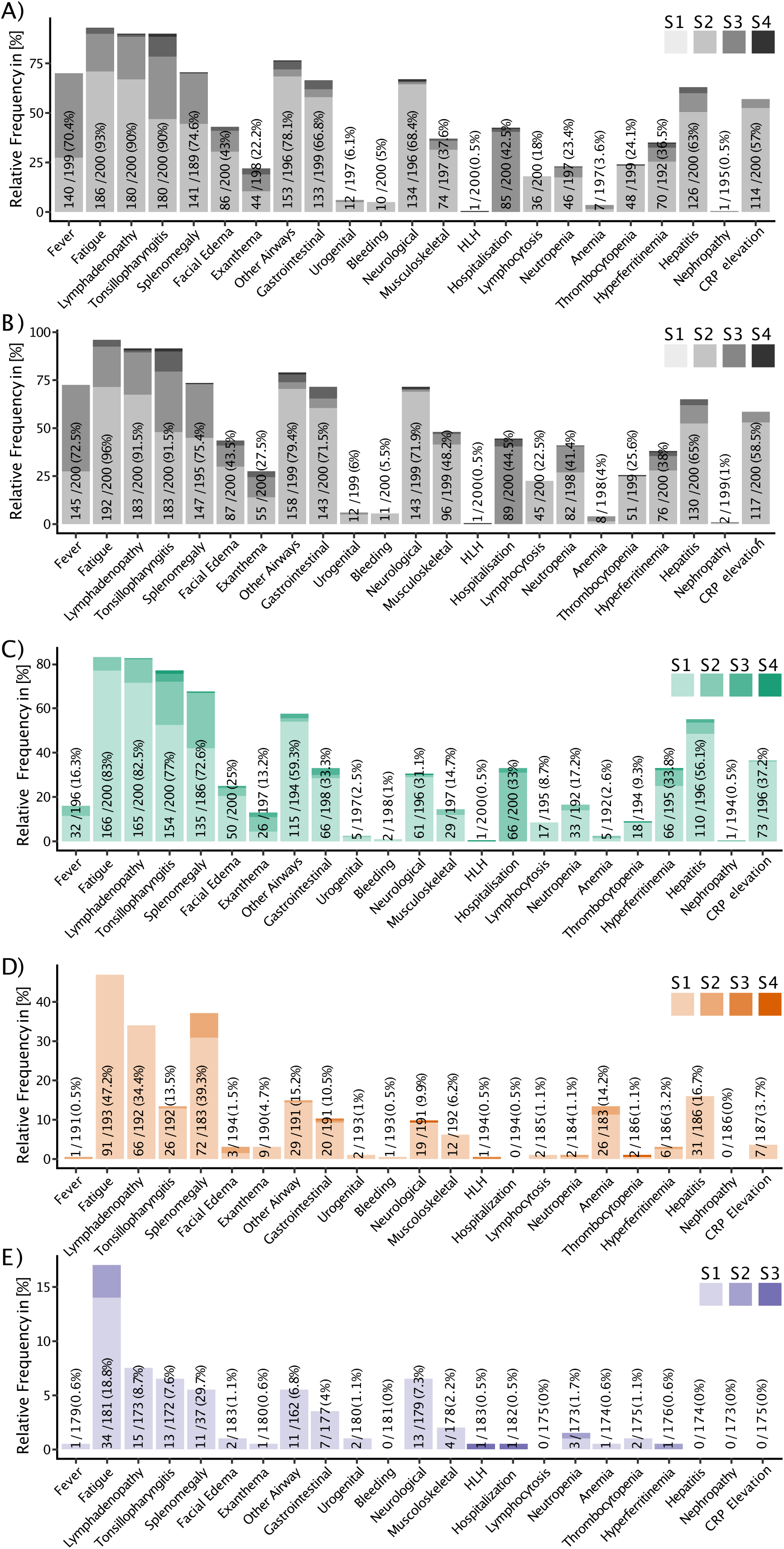
IMMUC Severity Scores of Each Symptom. The severity of individual symptoms for the very early period (VEP, A), the early period (EP, B), and at the visits V1 (C), V2 (D), and V3 (E) are shown. n/N (%) indicates the number of patients with the symptom relative to the number of patients where the respective symptom was assessed.

