## Supplementary Material M1 for "Predictors of Postviral Symptoms Following Epstein-Barr Virus-Associated Infectious Mononucleosis in Young People – Data from the IMMUC Study"

**Patient ID:** EP- ... ... ... ... - ... ... / ... ... ... - ... ... - ...

*If applicable add/ underline terms in brackets*

### ☐ Congenital immunodeficiency / warning signs

☐ no ☐ yes Known congenital immunodeficiency (…………………………………………………………………………………)
☐ no ☐ yes Known failure to thrive
☐ no ☐ yes ≥ 8 minor infections per year (common cold, tonsillitis, pharyngitis, otitis media, lymph node abscess on the neck, bronchitis, gastroenteritis)
☐ no ☐ yes Major infection (pneumonia, sepsis, meningitis, septic arthritis, osteomyelitis, empyema, deep abscess)
☐ no ☐ yes Unusual pathogens (mycosis, pseudomonas, others: ……………………………………………………………..)
☐ no ☐ yes Chronically-recurring infections (……….. ………………………………………………………………………)
☐ no ☐ yes Localization (……………………………………………………………………………………………………….)
☐ no ☐ yes Residuals of infections (bronchiectasis, scars, loss of nails, others: ……………………………………….)
☐ no ☐ yes Granuloma („sarcoid-like lesions“) (skin, bowels, other localization: ……………………………………...)
☐ no ☐ yes Autoimmune disease (thyroiditis, diabetes mellitus type I, rheumatoid arthritis, multiple sclerosis, systemic lupus erythematosus, immune cytopenia, others: ...……………………………………………......)
☐ no ☐ yes Recurrent fever (unclear genesis) (frequency: ...………………………………………………………...)
☐ no ☐ yes Unclear skin rash / eczema / neurodermatitis
☐ no ☐ yes Lymph proliferation (chron. lymphocytosis, chronic/ recurrent lymph node swelling, chron. splenomegaly)
☐ no ☐ yes Chronic bowel inflammation (specific diagnosis: ………………………………………...……………..........)

### ☐ Other pre-existing illnesses

☐ no ☐ yes Viral infection with known pathogen (influenza/ Influenza virus, herpes labialis/ herpes simplex virus [HSV], chickenpox/ herpes zoster/varicella-zoster virus [VZV], measles/ measles virus, three-day fever/ pityriasis rosea/ human herpesvirus 6 [HHV6]/ HHV7), others: ……………………………………………………………………………………………………………………...)
☐ no ☐ yes Bacterial infection with known pathogen or antibiotic therapy (scarlet fever/ group A streptococcus [GAS]), others: ……………………………………………………………………………………………………)
☐ no ☐ yes Febrile illness during the last 6 months, number of: ……………………………………………………...)
☐ no ☐ yes Infectious, non-febrile illness during the last 6 months, number of ……………………………………...)
☐ no ☐ yes Allergies (pollen/ grass, foods, animal hair, insect venom, medicine, others: ………………………………… ……………………………………………………………………………………………………………………)
☐ no ☐ yes Oncological disease (leukemia/ lymphoma, others: ...………………………………………………………..)
☐ no ☐ yes Transplantation (………...……………………………………………………………………………………..)
☐ no ☐ yes Developmental delay (motoric, mental:…………………………………………………………………….....)
☐ no ☐ yes Transfusion/blood products (which/reason: ……………….……………..………………………..…………)
☐ no ☐ yes Other pre-existing illnesses ☐ dialysis ☐ migraine
☐ others (which: ………………………..……………………………………….........)

### ☐ Medication during the last 6 months (*prior to* onset of IM symptoms)

☐ no ☐ yes ☐ Vitamine D
☐ Antibiotics (which: ……………………………………………………………………………………..)
☐ Immunosuppressants (which: …......……………………………………………………………………..)
☐ Analgesics (which: ……………………………...…………………………………………………………..)
☐ Steroids (which: ……………………………...…………………………………………………………..)
☐ Others (which: ……………………………...…………………………………………………………..)

### ☐ Vaccination

☐ no ☐ yes ☐ Standard (STIKO[1] recommendation) ☐ standard (incomplete) ☐ VZV (chickenpox)
☐ During the last 6 months (STIKO, influenza, tick-borne encephalitis [TBE], human papillomavirus [HPV], yellow fever, others……………………………………………………………………………………………………)
☐ Additional vaccinations (influenza, TBE, HPV, yellow fever, others: ......……………..……………….…..)

### ☐ Exposure to farm animals and/or pets

☐ no ☐ yes Farm animals
☐ no ☐ yes Pets (which: ………………………………………………………………………………………………..)

[1] “Ständige Impfkommission” – the standing committee on vaccination at the Robert Koch Institute (RKI), a department of the German Federal Health Agency

**Patient ID:** EP- ... ... ... ...

**☐ Travel outside of Germany during the last 6 months**

☐ no ☐ yes ☐ Europe/ Middle East:…………………..… ☐ Far East:……………….………………
☐ other:…………………………………..….

**☐ Family history**

☐ no ☐ yes Consanguineous parents (if yes, degree: ……………………………………………………………………………)
☐ no ☐ yes (Half)siblings total (how many:……………………………………………………………………………………..)
☐ no ☐ yes (Half)siblings in same household (how many: …………………………………………………………………...)
☐ no ☐ yes Persons in same household (how many: ………….. <25 years : ……………………………………………....)
☐ no ☐ yes Infectious Mononucleosis (how many:……………………...……………………………………………………....)
☐ no ☐ yes Congenital immunodeficiency (which/ who:…………..…………………………………………………………..)
☐ no ☐ yes Transplantation (which/ who:………………………………………………………………………………………)
☐ no ☐ yes Oncological disease (which/who/age (above/ below 50 years) ……………………………………………….
………………………………………………………………………………………………………………………
……………………………………………………………………………………………………………………..)

☐ no ☐ yes Autoimmune disease (thyroiditis, diabetes mellitus type I, rheumatoid arthritis, multiple sclerosis, systemic lupus erythematosus, immune cytopenia, others: ..……..……………………………………..……......................
……………………………………………………………………………………………………………………..)
☐ no ☐ yes Allergies (which/ who: ………………………………..…………………………………………………………...
……………………………………………………………………………………………………………………..)
☐ no ☐ yes Other pre-existing illnesses (which/ who: ……………………………..………..…………..……………………
……………………………………………………………………………………………………………………)
