## Supplementary Material M2 for "Predictors of Postviral Symptoms Following Epstein-Barr Virus-Associated Infectious Mononucleosis in Young People – Data from the IMMUC Study"

IMMUC Score ____________________________________________

### IMMUC Score – Symptom and Severity Score

| **Patient ID:** EP- … … … … - … … / … … … - … … - … | | **IMMUC Score** |
|---|---|---|
| **Total Complexity** | *Number of clinical and laboratory symptoms* | |
| **Clinical Complexity** | *Number of clinical symptoms* | |
| **Laboratory Complexity** | *Number of laboratory symptoms* | |
| **Severity of all symptoms** | *Maximum severity score of clinical and laboratory symptoms; (s5 – death due to IM)* | |
| **Severity of Clinical Symptoms** | *Maximum severity score of clinical symptoms* | |
| **Severity of laboratory Symptoms** | *Maximum severity score of laboratory symptoms* | |

### Clinical Symptoms

| **Symptom** | **Severity** |
|---|---|
| **1. Fever** | |
| No | **s0** |
| ≥ 38.5 – 39.4°C | **s1** |
| ≥ 39.5°C | **s2** |
| Antibiotics | |
| No information | **sx** |
| **2. Fatigue** | |
| No | **s0** |
| Can walk alone | **s1** |
| Assisted walking | **s2** |
| T6: Reduced education/work/ club sports | |
| Can not walk | **s3** |
| No information | **sx** |
| **3. Lymphadenopathy** | |
| No | **s0** |
| Enlarged nodes | **s1** |
| Painful nodes | |
| Movement locally impaired | **s2** |
| Antibiotics | |
| Computed tomography (CT)/ Magnetic resonance imaging (MRI) | **s3** |
| Biopsy (Bone marrow biopsy [BMB]/ Lymph node biopsy [LNB]) | **s4** |
| No information | **sx** |
| **4. Tonsillopharyngitis** | |
| No | **s0** |
| Sore throat, pain killers | **s1** |
| Snoring/upper respiratory tract obstruction | |
| Tonsillitis | |
| Antibiotics | **s2** |
| Steroids | **s3** |
| Oxygen supply | |
| Tonsillotomy | **s4** |
| No information | **sx** |
| **5.Splenomegaly** | |
| No | **s0** |

IMMUC Score ___________________________________________

| | |
|---|---|
| Sonography >100-119%, not palpable | **s1** |
| Palpable | |
| Sonography >120% | **s2** |
| Rupture | **s4** |
| Surgery | |
| No information | **sx** |
| **6. Facial edema** | |
| No | **s0** |
| No visual impairment | **s1** |
| Partial visual impairment | **s2** |
| Antihistamines | |
| Antibiotics due to lid phlegmon | |
| Complete visual impairment | **s3** |
| Steroids | |
| No information | **sx** |
| **7. Exanthema** | |
| No | **s0** |
| Maculopapulous w/o Aminopenicillin | **s1** |
| Maculopapulous post Aminopenicillin | |
| Aphthae | |
| Itching a/o pain | **s2** |
| Antihistamines | |
| Local steroids | |
| Erythema nodosum | |
| Bullae | **s3** |
| Steroids | |
| Biopsy | |
| Necrosis | **s4** |
| No information | **sx** |
| **08. Bleeding, Treatment of bleeding or anemia** | |
| No | **s0** |
| Nose bleeding (≤10min) | **s1** |
| Hematoma (≤10cm) | |
| Petechiae | |
| Hematoma (>10cm) | **s2** |
| Cyclocaproic acid as anticoagulans | |
| nose bleeding (>10min a/o >3x/d) | **s3** |
| Oral blood bullae | |
| Steroids | |
| Evans syndrome | **s4** |
| Organ bleeding | |
| Rituximab | |
| Transfusion | |
| Plasmapheresis | |
| No information | **sx** |
| **09. Other airways** | |
| No | **s0** |
| Otitis | **s1** |
| Rhinitis | |
| Hoarseness | |
| Cough | |
| Stress-related mild dyspnoe | |
| Parotitis | |
| Swollen a/o tender tongue | |
| Antibiotics | **s2** |
| Local steroids | |
| Oxygen supply | **s3** |
| Steroids | |
| X-ray thorax | |
| Pneumonia | |
| CT thorax | **s4** |
| Paracentesis/ ear ventilation tube/ adenoidectomy | |
| Thoracic drainage | |
| Mechanical ventilation | |
| No information | **sx** |

IMMUC Score ________________________________________________

| 10. Other gastrointestinal | |
|---|---|
| No | **s0** |
| Abdominal pain | **s1** |
| Nausea, vomiting | |
| Watery diarrhea | **s2** |
| Melaena a/o Hematemesis | |
| Antiemetics | |
| Weight loss >10%, possibly not only Epstein-Barr virus (EBV) | |
| Weight loss >10% | **s3** |
| Acute abdomen, pancreatitis, appendicitis, ileus | |
| Abdominal X-ray, CT/MRI | |
| Antibiotics | |
| Endoscopy | **s4** |
| Surgery | |
| No information | **sx** |
| **11. Urogenital** | |
| No | **s0** |
| Dysuria | **s1** |
| Genital ulcer | **s2** |
| Gross hematuria | |
| Antibiotics | |
| Nephrotic syndrome | **s3** |
| Nephritis | |
| Nephrological drug treatment | |
| Dialysis | **s4** |
| Renal biopsy | |
| No information | **sx** |
| **12. Neurological** | |
| No | **s0** |
| Headache | **s1** |
| Meningism | **s2** |
| Lumbar puncture | |
| Antibiotics | |
| Seizure | **s3** |
| Documented meningitis | |
| Neurological drug 1x | |
| CT/MRI | |
| Electroencephalogram (EEG), nerve conduction velocity (NCV), Electrophysiology (EP)i | |
| Neurological drug > 1x | **s4** |
| Encephalitis | |
| Neuritis | |
| No information | **sx** |
| **13. Musculoskeletal** | |
| No | **s0** |
| Body aches | **s1** |
| Painkillers for body aches | **s2** |
| Joint swelling | |
| Joint ultrasound | **s3** |
| Pain-related immobilisation | |
| No information | **sx** |
| **14. Imminent or treatment for hemophagocytic lymphohistiocytosis (HLH)** | |
| No | **s0** |
| 4 of 8 criteria fulfilled | **s3** |
| Steroids | |
| Other HLH drug(s) | **s4** |
| Stem cell Tx | |
| No information | **sx** |
| **15. Hospitalization** | |
| No | **s0** |
| Normal ward < 8d | **s2** |
| Normal ward ≥8d a/o intermediate care unit | **s3** |
| Intensive care unit (ICU) | **s4** |
| No information | **sx** |

IMMUC Score ______________________________________________

### Laboratory Symptoms

| | |
|---|---|
| **16. Lymphocytosis** | |
| No | **s0** |
| ≥ 1,5x normal (%) | **s1** |
| No information | **sx** |
| **17. Neutropenia** | |
| No | **s0** |
| 1.0 - 1.49 G/l | **s1** |
| 0.5 - 0.99 G/l | **s2** |
| < 0.5 G/l | **s3** |
| No information | **sx** |
| **18. Anemia** | |
| No | **s0** |
| 9.0- 10.0 g/dl | **s1** |
| 7.0 – 8.9 g/dl | **s2** |
| 5.0 – 6.9 g/dl | **s3** |
| < 5 g/dl | **s4** |
| Reticulocyte production index >3 | |
| Reticulocyte production index <2 | |
| No information | **sx** |
| **19. Thrombocytopenia** | |
| No | **s0** |
| 50 – 149 G/l | **s1** |
| 30 – 49 G/l | **s2** |
| 10 – 29 G/l | **s3** |
| < 10 G/l | **s4** |
| No information | **sx** |
| **20. Hyperferritinemia** | |
| No | **s0** |
| ≥ 100 ug/l > norm | **s1** |
| ≥ 500 – 999 ug/l | **s2** |
| ≥ 1000- 4999 ug/l | **s3** |
| ≥ 5000 ug/l | **s4** |
| No information | **sx** |
| **21. Hepatitis, Cholangitis** | |
| No | **s0** |
| Alanine aminotransferase (ALAT) u/o aspartate aminotransferase (ASAT) 2,0 - 9,9 x normal | **s1** |
| ALAT u/o ASAT 10,0 - 19,9 x normal | **s2** |
| ALAT u/o ASAT ≥20,0 x normal | **s3** |
| Ammonia ↑, Cholinesterase↓ u/o Quick value < 50% | **s4** |
| No information | **sx** |
| **22. Nephropathy** | |
| No | **s0** |
| Creatinine ↑ ≤ 0,2 mg/dl over norm + elevated Cystatin C | **s1** |
| Creatinine ↑ > 0,2 mg/dl over norm, but < 1,5 mg/dl (<10a) + elevated Cystatin C | **s2** |
| Creatinine ↑≥ 0,2 mg/dl over norm, but < 2,0 mg/dl (≥10a) + elevated Cystatin C | |
| Creatinine ↑ ≥ 1,5 mg/dl (<10a) + elevated Cystatin C | **s3** |
| Creatinine a ≥ 2,0 mg/dl (≥10a) + elevated Cystatin C | |
| No information | **sx** |
| **23. C-reactive protein (CRP) elevation** | |
| No | **s0** |
| > 10 – 99 mg/l | **s1** |
| 100 – 199 mg/l | **s2** |
| Antibiotics | |
| ≥ 200 mg/l | **s3** |
| No information | **sx** |
